# Dynamic phospho-proteogenomic analysis of gastric cancer cells suggests host immunity provides survival benefit

**DOI:** 10.1101/2024.02.06.24302407

**Authors:** Kohei Kume, Midori Iida, Takeshi Iwaya, Akiko Yashima-Abo, Yuka Koizumi, Akari Konta, Kaitlin Wade, Hayato Hiraki, Valerie Calvert, Julia Wulfkuhle, Virginia Espina, Doris R. Siwak, Yiling Lu, Kazuhiro Takemoto, Yutaka Suzuki, Yasushi Sasaki, Takashi Tokino, Emanuel Petricoin, Lance A. Liotta, Gordon B. Mills, Satoshi S. Nishizuka

## Abstract

The mainstay of advanced gastric cancer (GC) therapy is DNA-damaging drugs. Using proteogenomic analysis of a panel of eight GC cell lines, we identified genetic alterations and signaling pathways, potentially associated with resistance to DNA-damaging drugs. Notably, 5-fluorouracil (5FU) resistance was associated with PD-L1 expression, but not established GC subtypes. In publicly available cohort data, PD-L1 expression was associated with a reduced risk of GC progression. In addition to PD-L1, expression of inflammatory genes induced by lymphocyte cytokines was consistently associated with prolonged survival in GC. In our validation cohort, total lymphocyte count (TLC) predicted a better relapse-free survival rate in GC patients with 5FU-based adjuvant chemotherapy than those with surgery alone. Moreover, TLC^+^ patients who had no survival benefit from adjuvant chemotherapy were discriminated by IκBα expression. Collectively, our results suggest that 5FU resistance observed in cell lines may be overcome by host immunity or by combination therapy with immune checkpoint blockade.

## Introduction

Advanced gastric cancer (GC) is a leading causes of cancer death worldwide and there are limited treatment strategies for patients with this cancer^1^. Multidisciplinary treatments, including combinations of surgery, chemotherapy, and radiotherapy have been used to improve survival for patients with GC^2^. Indeed, curative surgery followed by adjuvant chemotherapy with an oral fluoropyrimidine S-1 containing the 5-fluorouracil (5FU) prodrug tegafur^3^ decreases relapse and extends disease-free survival, particularly in the Japanese population^4,5,6^. Unfortunately, a substantial number of patients still experience relapse. In the Japanese population, most relapses occur after S-1 adjuvant chemotherapy, suggesting that acquired 5FU-resistance likely plays a substantial role for relapse. Clarifying the mechanisms leading to 5FU-resistance offers the opportunity to select molecular targeting drugs designed to prevent relapse after treatment with DNA-damaging drugs.

Most GCs arise from glandular epithelia of the gastric mucosa that is exposed to a range of substances and stimulants that can promote host immune and inflammatory responses. Excessive activation of nuclear factor-κB (NFκB), which is considered to be a hallmark of inflammation-associated cancers^7^, has been demonstrated to play a crucial role in GC progression and relapse^8,9,10^. NFκB is activated not only by cytokines but also DNA-damaging drugs via the cytosolic DNA-sensing pathway^11^. Stimulator of interferon genes (STING) is a key component of this DNA-sensing pathway and engages an IκB kinase (IKK) complex that phosphorylates inhibitor of nuclear factor-κB (IκB) proteins to target them for proteasomal degradation. This degradation of IκB proteins allows NFκB/Rel transcription factors to enter the nucleus with subsequent activation of genes related to proinflammatory signaling. Despite the strong rationale for this pathway, there is limited information concerning the functional consequences of NFκB-mediated proinflammatory signaling in the response of GCs to 5FU.

Cancer cell lines are important model systems to study quantitative cellular and molecular response to external stimuli and drug uptake. 5FU-resistant GC cell lines have been used to demonstrate specific roles for 5FU metabolism, prostaglandin production, and autophagy in 5FU resistance^12,13,14^. The cell lines MKN45/5FU (45FU) and MKN74/5FU (74FU) were established from the poorly differentiated MKN45 and differentiated MKN74 cell lines, respectively, by culturing in the presence of increasing concentrations of 5FU over the course of one year^15^. Similar to previous work on tissue samples from 5FU-nonresponders^16^, these cell lines showed increased expression of *thymidylate synthase (TYMS)* and decreased expression of *orotate phosphoribosyltransferase (OPRT)* compared to their parental counterparts. However, comprehensive knowledge of their genomic, transcriptomic, and proteomic profiles associated with phenotypic drug response to GC treatment is limited. Although the Cancer Genome Atlas (TCGA) and the Asian Cancer Research Group (ACRG) provide comprehensive genomic and transcriptomic profiles of GCs and further established a robust GC molecular classification method^17,18^, phenotypic drug responses and proteomic profiles have not yet been integrated.

In the present study, we used our dynamic phosphoproteomics platform, reverse phase protein array (RPPA), with GC cell lines to determine the association between key molecules that have been validated in real-world cohort data and the host immune responses. Our findings suggest that the mechanisms of 5FU resistance observed in cell line models can be counteracted by lymphocyte-mediated host immunity potentially in the presence of immune checkpoint blockade.

## Results

### Comprehensive proteogenomic profiles of GC cell lines with phenotypic drug response

We started by characterizing GC cell lines in terms of fifty percent growth inhibitory concentration (GI_50_) for different chemotherapeutic agents, GC-associated mutations^17,18,19^, and RNA and protein expression profiles (**Fig. 1**). Although 5FU resistance was observed for both 45FU and 74FU cell lines^15^, 74FU also had cisplatin (CIS) and etoposide (ETP) resistance, indicating cross resistance among DNA-damaging drugs (**Fig. 1A**; **Supplementary Fig. 1A** and **1B**). Neither cell line showed apparent resistance to the non-DNA damaging drug docetaxel (DTX) that function as a mitotic inhibitor. As expected from gene alterations reported for GC, all cell lines tested carried *TP53* mutations (**Fig. 1B**). The cell line MKN1 carried a *PIK3CA* mutation, one of the key determinants of the Epstein-Barr virus (EBV)-positive GC subtype (**Fig. 1B**), but none of the eight cell lines tested showed EBV gene expression (**Supplementary Fig. 1C**). Copy number variation (CNV) analysis revealed a positive correlation between the number of CNV loci and CIS sensitivity (**Fig. 1C**). This correlation resembles CIS hypersensitivity of cells derived from Fanconi anemia patients as a phenotypic consequence of genomic instability^20^. CIS sensitivity was also associated with a lack of vimentin silencing^21,22^ (**Fig. 1D**). As reflected by their differential drug responses, 45FU and 74FU apparently do not share similarly expressed genes and proteins, except for *CDH1*, which encodes E-cadherin (**Fig. 1E**). Indeed, both lines retained most of the genetic programs of their parental cells (**Supplementary Fig. 1D**). Thus, we further examined reasons for the correlation between CIS sensitivity and increased number of CNV loci, and then pursued unbiased RPPA analysis to identify shared signaling outcomes between 45FU and 74FU.

**Figure 1.**
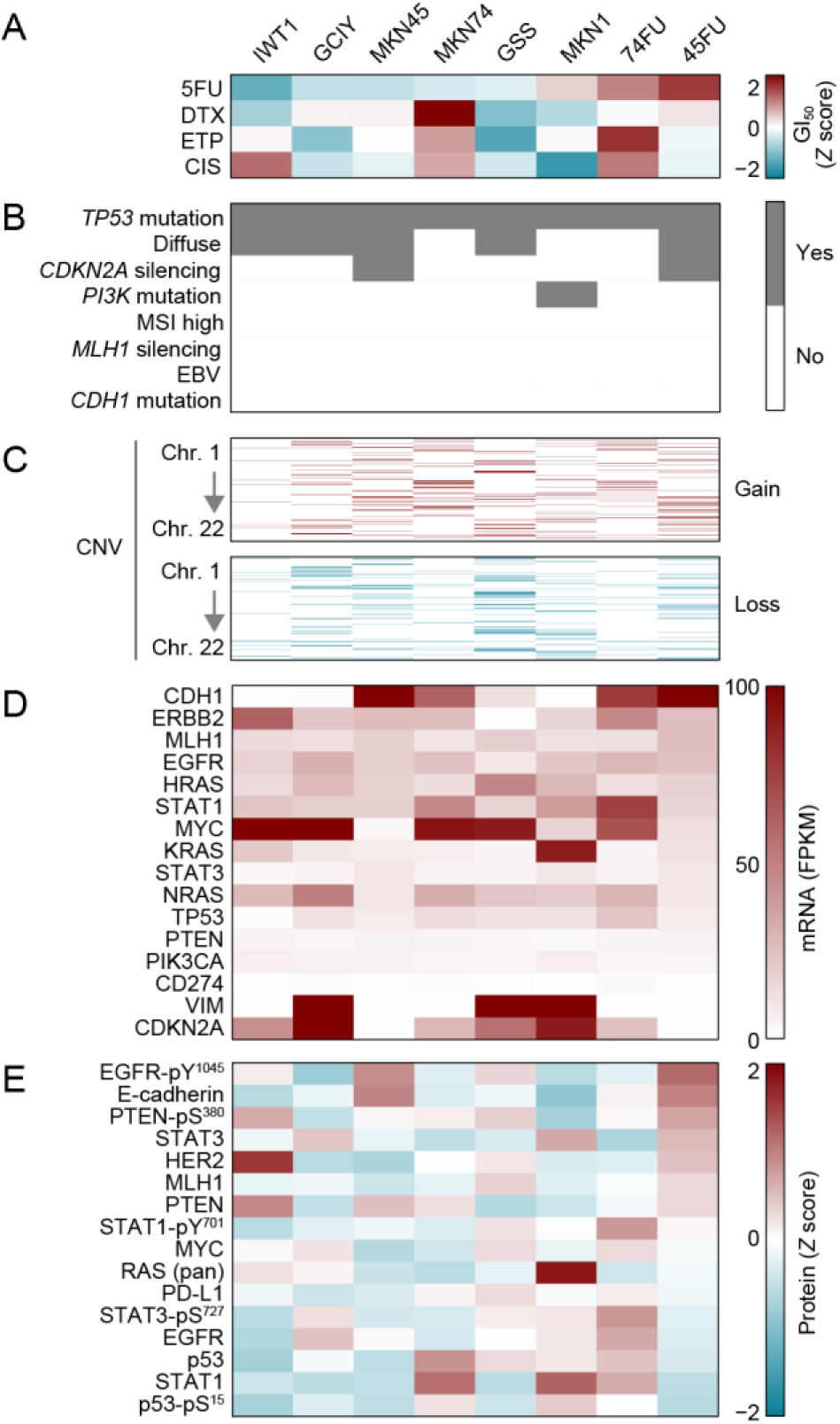
Integrative multiplatform analyses of GC cell lines. (**A**−**E**) GI_50_ heatmap for each drug tested (**A**), cell line summary (**B**), copy number variation (CNV) landscape (**C**), and mRNA and protein expression heatmaps (**D** and **E**) are depicted. Cell lines are ordered by CIS sensitivity. Relevant genes for cell line summary and expression profiles were selected based on previous studies of GC subtypes^17,18^.

### CIS-sensitive GC cells predominantly demonstrate CIN

To examine reasons for the correlation between CIS sensitivity and increased number of CNV loci, we segregated CIS-sensitive and -resistant cell lines based on their GI_50_ CIS profiles (**Fig. 2A**). As expected from Fig. 1, the same cell lines showed CIS and ETP sensitivity and resistance. This observation is further supported by the PRISM repurposing primary screen^23^, showing a positive correlation between CIS and ETP sensitivity for both gastric and colon cancer cell lines (**Fig. 2B** and **2C; Supplementary Table 1**). Although no difference in CNV gain is observed, a 3-fold higher level of CNV loss was detected in dual CIS/ETP-sensitive cell lines compared to their resistant counterparts (**Fig. 2D**). Genes that were frequently lost included *c-kit* (*KIT*), *kinase insert domain-containing receptor* (*KDR/VEGFR2*), and *adhesion G protein-coupled receptor L3* (*ADGRL3*), which are all located on chromosome at 4q12–q13 (**Fig. 2E**). Moreover, scores for CIS and ETP sensitivity for gastrointestinal cell lines from the PRISM repurposing primary screen were negatively correlated with *KIT* copy number (**Fig. 2F**). Thus, CIS sensitivity in GC cell lines could be defined by chromosomal instability (CIN), represented by loss of proximal 4q12–q13, which could be related to the *KIT* copy number.

**Figure 2.**
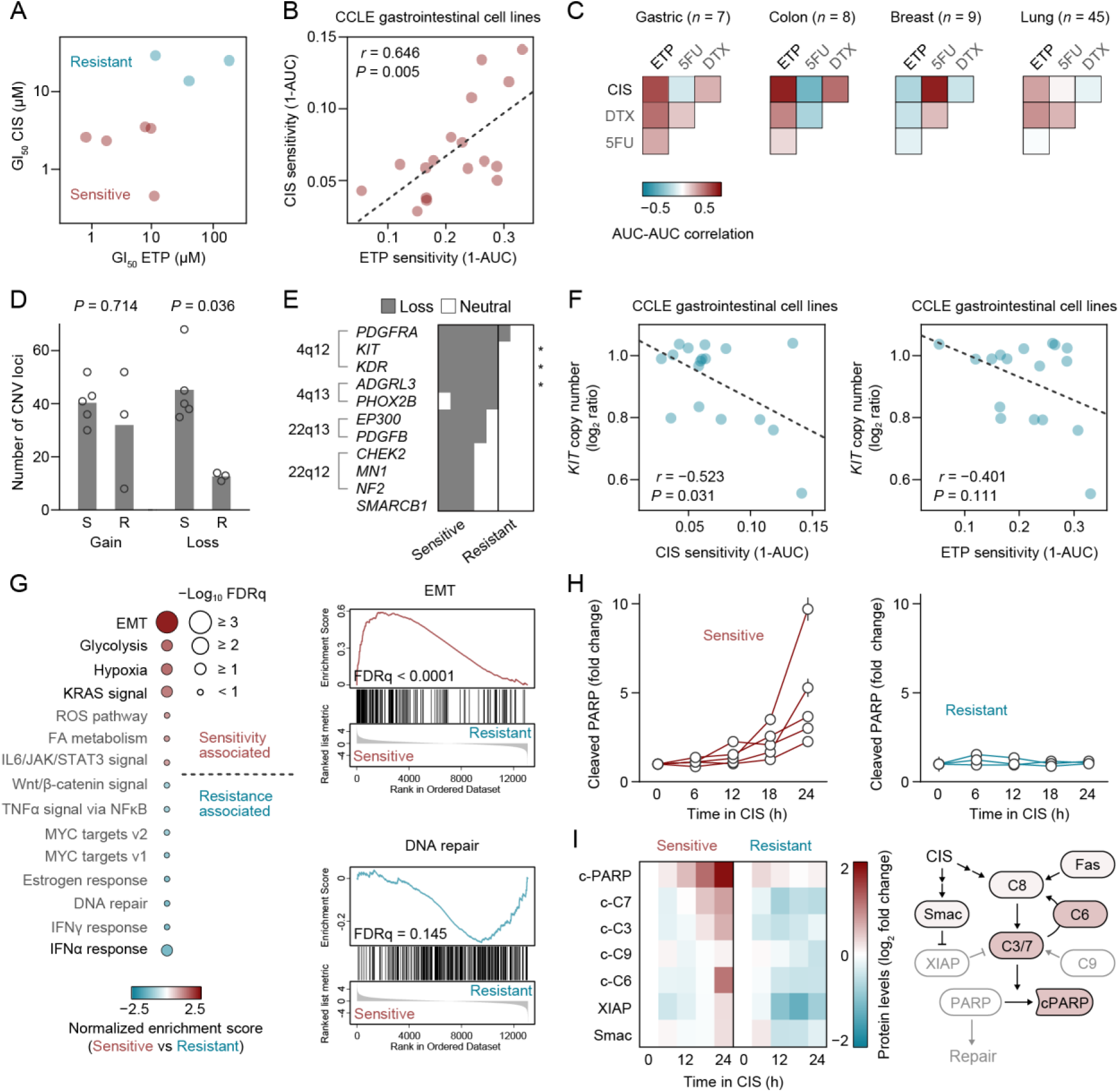
CNV and mesenchymal gene expression in CIS-sensitive GC cells. (**A**) GI_50_ profiles of GC cell lines to define sensitivity and resistance to CIS and ETP. (**B**) Scatter plot showing correlation between area under the dose-response curve (AUC) values for CIS and ETP in gastrointestinal cancer cell lines (right). Each dot indicates an individual cell line (*n* = 17) shared between the AUC data sets. (**C**) Heatmaps represent pairwise Pearson correlations between AUC values of CIS and ETP assessed in the PRISM repurposing secondary screen^23^. (**D**) Number of CNV loci detected. Each dot indicates an individual cell line. The two-tailed *P* values were obtained with a nonparametric Mann-Whitney *U* test. (**E**) Binary matrix representing genes that commonly lost its copy number in cell lines with dual CIS/ETP sensitivity. The two-tailed *P* values were obtained with Fisher’s exact test. (**F**) Scatter plot showing correlation between AUC values of CIS or ETP and *KIT* copy number in CCLE gastrointestinal cancer cell lines. Each dot indicates an individual cell line. (**G**) Hallmark GSEA signatures from RNA-seq data ranked by Normalized Enrichment Score (NES) for CIS/ETP-sensitive vs. CIS/ETP-resistant cell lines (left). GSEA plots showing positive and negative enrichment of “EMT” and “DNA repair” gene sets in CIS/ETP-sensitive cell lines (right). FDRq, false discovery rate (*q* value). (**H**) Time course RPPA data showing changes in cleaved PARP levels after CIS treatment. Error bars represent s.e.m. (**I**) Caspase-focused RPPA analysis of dual CIS/ETP-sensitive (mean, *n* = 5) and -resistant (mean, *n* = 3) cell lines (left) and a schematic of CIS-activated signaling outcomes in dual CIS/ETP-sensitive cell lines (right). c-PARP, cleaved PARP; c-C3−9, cleaved caspase-3−9.

### Lack of vimentin silencing is associated with CIS/ETP sensitivity induced by CIN

We next investigated the positive correlation between CIS sensitivity and the lack of vimentin silencing. As expected, gene set enrichment analysis (GSEA) showed that “epithelial-mesenchymal transition (EMT)” was the most enriched upregulated gene set in cell lines having dual CIS/ETP-sensitivity (**Fig. 2G**, left). RNA-seq gene expression and mass spectrometry-based proteomic data from the Cancer Cell Line Encyclopedia (CCLE) also showed a positive correlation between both vimentin gene and protein expression with CIS sensitivity across gastrointestinal cell lines (**Supplementary Fig. S2A** and **S2B**), which complements our RPPA-based proteomic data. TWIST, one of the master transcription factors of the vimentin gene, is directly regulated by hypoxia-inducible factor 1α (HIF1A), a key regulator of glycolytic genes, and promotes metastasis^24^. Supporting this transcriptional mechanism, “Glycolysis” and “Hypoxia” genes are concurrently enriched for cell lines having dual CIS/ETP sensitivity (**Fig. 2G**, left). These cell lines also showed negative enrichment of “interferon (IFN)α response” and “IFNγ response” pathways. Interestingly, recent work showed that CIN-induced chronic activation of cyclic GMP– AMP synthase (cGAS)-STING, which is a driver of cancer metastasis, led to STING depletion, thereby reducing IFN responsiveness^25^. Therefore, the lack of vimentin silencing in dual CIS/ETP-sensitive cell lines could be one of the consequences of chronic cGAS-STING activation induced by CIN.

### CIS/ETP-induced cell death pathway shares caspase effectors

We also explored central effectors of cell death pathways that are activated by CIS and ETP in GC cells. Consistent with the correlation between CIS/ETP sensitivity and CIN, downregulated genes are significantly enriched for the “DNA repair” pathway (**Fig. 2G**). Pairwise comparisons of time course protein expression profiles between dual CIS/ETP-sensitive and -resistant cell lines revealed consistently low levels of cleaved PARP in dual CIS/ETP-resistant cell lines regardless of CIS or ETP treatments (**Fig. 2H**; **Supplementary Fig. S2C**). Interestingly, *PARP*^−/−^ mice were reported to exhibit chromosomal aberrations, including gain and loss in regions of chromosomes 4, 5, and 14^26^. PARP is a well-established target of caspase cleavage. Subsequent caspase-focused analysis highlighted lack of cleaved caspase-3, -7, and -6 in dual CIS/ETP-resistant cell lines following CIS treatment (**Fig. 2I**). Both cleaved caspase-3 and -7 cleave PARP to inhibit its enzymatic activity^27^, whereas cleaved caspase-6 creates a positive feedback loop with caspase-8, -7, and -3^28,29^. Therefore, caspase-6 cleavage appears to be a key event in the CIS response in GC cells. Meanwhile, levels of cleaved caspase-9 and X-linked inhibitor of apoptosis protein (XIAP) concurrently increased in response to ETP treatment, reflecting the early plateau of PARP cleavage (**Supplementary Fig. S2D**). Together, these results suggest that caspase-3 and -7 are the central effectors of cell death pathways activated by CIS and ETP in GC cells.

### 5FU-induced MAPK signaling pathways converge on phosphorylation of MEK1

Next, we pursued proteogenomic analysis to identify shared signaling outcomes in the two 5FU-resistant cell lines, 45FU and 74FU (**Fig. 3A**). Pairwise comparison of targeted pharmacogenomic and comprehensive cancer panel sequencing-based mutation profiles between 5FU-resistant and matched parental cell lines identified a missense mutation in *neurofibromatosis type 1* (*NF1*), c.5461G>T (V1821F) that is unique to the 45FU cell line (**Fig. 3B** and **3C**). *NF1* encodes a GTPase-activating protein (GAP) that negatively regulates Ras pathway activity by accelerating the hydrolysis of Ras-bound GTP, thereby acting as a tumor suppressor^30^. However, *NF1*^V1182F^ is not listed in the Genome Aggregation Database (gnomAD) (https://gnomad.broadinstitute.org/) as a pathogenic mutation. Previous studies showed that a missense mutation in *dihydropyrimidine dehydrogenase* (*DPYD*), c.85T>C (C29R), confers gain-of-function activity^31,32^. Despite the three-to five-fold difference in 5FU sensitivity between the 5FU-resistant and matched parental cell lines, both pairs harbor the same *DPYD*^C29R^ mutation. Consistent with previous reports^15,16^, both 5FU-resistant cell lines showed a two-to four-fold increase in *TYMS* expression compared to their matched parental cell lines in the presence of 5FU, suggesting that enhanced 5FU metabolism contributes in part to the phenotypic 5FU response (**Supplementary Fig. 3A**). Although we detected no unique mutations in 74FU cells, the parental cell line MKN74 has a unique mutation in *fibroblast growth factor receptor 2* (*FGFR2*), c.1199G>A (R400Q), suggesting that this cell line contains a minor subpopulation with wild-type *FGFR2* that confers a survival advantage in the presence of 5FU. In contrast, 74FU cells harbor a missense mutation in *Tet methylcytosine dioxygenase 2* (*TET2*), c.367C>T (R123C), which was not detected in the parental MKN74 cells. Unlike dual CIS/ETP sensitivity determined by the loss of proximal 4q12–q13, pairwise comparison of CNV profiles between the 5FU-resistant and matched parental cell lines identified no copy number changes shared by 45FU and 74FU cell lines (**Fig. 3D**). To identify proteins associated with 5FU resistance, we thus determined the correlation between protein level and GI_50_ 5FU using Spearman’s rank correlation analyses (**Fig. 3E**). At baseline, signal transducer and activator of transcription (STAT1)-pY^701^, MAP-extracellular signal-regulated kinase (MEK) 1-pS^298^, and Rac1 p21-activated kinase (PAK) 1-pT^423^ are the top three proteins that positively correlated with GI_50_ 5FU. At 24 h after 5FU treatment, MEK1-pS^298^, but not STAT1-pY^701^ and PAK1-pT^423^, remained as highly expressed, suggesting that MEK1-pS^298^ is a key driver molecule of 5FU resistance.

**Figure 3.**
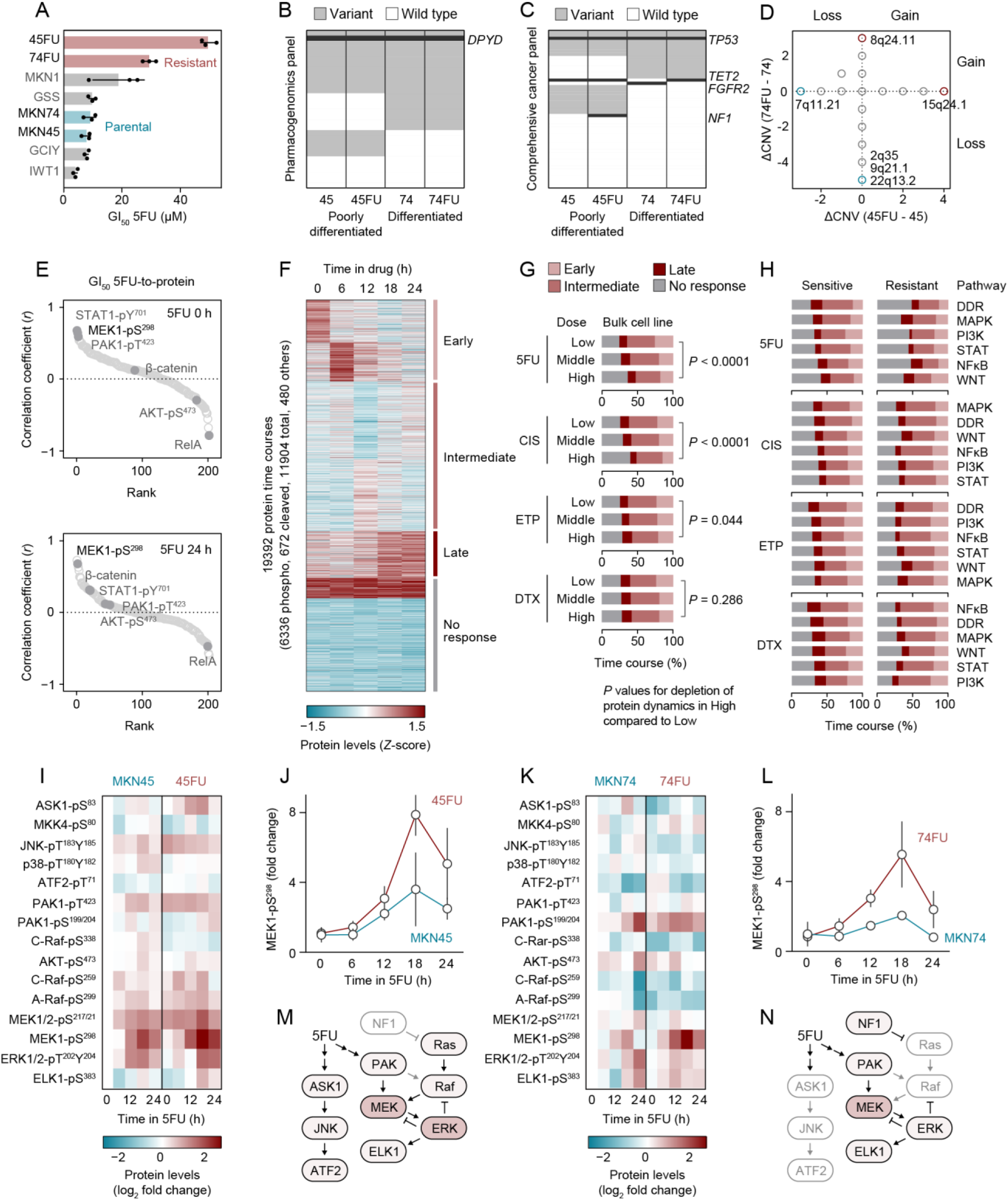
5FU-induced MAPK signaling pathways. (**A**) GI_50_ profiles of GC cell lines to define 5FU sensitivity and resistance. Two pairs of 5FU resistant and parental cell lines are highlighted. (**B**) Pharmacogenomics panel shows drug metabolizing genes harboring single-nucleotide variants (SNVs) in each cell line. *DPYD* is highlighted as a canonical 5FU metabolism pathway gene. (**C**) Comprehensive cancer panel. *TP53*, *TET2*, *FGFR2*, and *NF1* are highlighted for common, MKN74-unique, and 45FU-unique mutations. (**D**) CNV analysis showing no shared copy number changes between 74FU and 45FU. Copy number changes were calculated by subtracting the copy number of each chromosomal position in matched parental cell lines from that in resistant cell lines (ΔCNV). Chromosomal positions with maximum and minimum ΔCNVs are highlighted. (**E**) Spearman’s rank correlation analysis to determine the correlation between GI_50_ 5FU and protein expression at baseline (top panel) and 24 h after 5FU treatment (bottom panel). (**F**) Temporal proteomic changes in eight cell lines within 24h of 5FU, CIS, ETP, or DTX treatment. Seven clusters were determined by K-means clustering and further grouped for early, intermediate, late, and no response based on their kinetics. (**G**) Proportions of protein expression time courses segregated into three doses for each drug. Two-tailed *P* values were obtained with Fisher’s exact test. (**H**) Proportions of protein expression time courses from high-dose conditions segregated into six signaling pathways including DNA damage response (DDR), MAPK, PI3K, STAT, NFκB, and WNT pathways. (**I**) MAPK pathway-focused RPPA analysis of 45FU and parental MKN45 cells. (**J**) Temporal changes in MEK1-pS^298^ levels after 5FU treatment. (**K**) MAPK pathway-focused RPPA analysis of 74FU and parental MKN74 cells. (**L**) Temporal changes in MEK1-pS^298^ levels after 5FU treatment. (**M**) 5FU-activated MAPK signaling pathways in 45FU cells. (**N**) 5FU-activated MAPK signaling pathways in 74FU cells. Error bars represent s.d. (**A**) or s.e.m. (**J** and **L**).

### Dynamic phosphoproteomics identifies MEK1 as a key 5FU-resistance pathway molecule

To see how 5FU affects signaling pathway activity in 5FU-resistant and -sensitive cell lines, we divided the protein expression time courses into four groups, early (peaked by 6 h), intermediate (peaked by 12 h), late (peaked by 24 h), and no response (flat), and then determined the proportions of these dynamics in different signaling pathways including DNA damage response (DDR), mitogen-activated protein kinase (MAPK), phosphoinositide-3-kinase (PI3K), STAT, NFκB, and WNT (**Fig. 3F–3H**). Compared to a non-DNA damaging drug DTX, DNA-damaging drugs including 5FU, CIS, and ETP reduced the proportions of early, intermediate, and late protein changes in a dose-dependent manner, indicating depletion of protein dynamics (**Fig. 3G**). The segregation of protein expression time courses by sensitive and resistant cell lines revealed that 5FU-resistant cell lines had lower changes in global protein expression dynamics compared to their sensitive counterparts, whereas the cell lines that are resistant to other drugs showed increased protein expression dynamics in response to each drug (**Fig. 3H**). Among the signaling pathways tested, the MAPK pathway appears to remain active in the 5FU-resistant cells having global protein dynamics depletion (**Fig. 3H**), which is consistent with the positive correlation between MEK1-pS^298^ levels and GI_50_ 5FU (**Fig. 3E**).

To identify MAPK signaling outcomes that are shared between 45FU and 74FU, we examined a phosphoproteomic analysis involving RPPA that was focused on the MAPK pathway. Among the three major MAPKs, c-Jun N-terminal kinase (JNK), p38, and extracellular signal-regulated kinase (ERK), JNK and p38 are activated by apoptosis signal-regulating kinase 1 (ASK1) in response to a diverse array of stresses such as oxidative stress, endoplasmic reticulum stress and calcium influx^33^, but no established stress-responsive upstream kinases have been reported for ERK. Interestingly, a previous study suggested that Rac1 GTPase, a PAK activator, plays an essential role in activation of gamma-irradiation-induced ERK1/2 signaling in the breast cancer cell line MCF7^34^. PAK has been demonstrated to phosphorylate S298 of MEK1 and S338 of C-Raf^35,36^. Therefore, we included PAK as an upstream kinase of Raf and MEK that can respond to 5FU. As expected from the genetic profiles, the effects of 5FU on signaling pathways differed between 45FU and 74FU cells (**Fig. 3I–3L**). The 45FU cells showed higher amplitude of ASK1 phosphorylation following 5FU treatment than did the parental cells, whereas phosphorylation levels of both ASK1 and its downstream kinase JNK were largely similar in both 74FU and its parental cell line (**Fig. 3I** and **3K**). In addition to ASK1, phosphorylation of A-Raf at S299 also increased in 45FU cells in response to 5FU, with baseline activity also being higher than in parental cells, potentially due to the loss-of-function *NF1* mutation in the cell line (**Fig. 3C**). Meanwhile, phosphorylation of C-Raf at S338 was instead depleted in 45FU cells, along with an increase in inhibitory phosphorylation at S259 (**Fig. 3I**). For 74FU cells, smaller or no changes in A-Raf phosphorylation were seen, and only phosphorylation of PAK being observed among kinases upstream of MEK (**Fig. 3K**). Notably, both 45FU and 74FU cells showed a sharp increase in MEK1 phosphorylation at S298 in response to 5FU (**Fig. 3J** and **3L**). However, phosphorylation of the downstream kinase ERK did not reflect the amplitude and timing of MEK1-pS^298^ peaks (**Supplementary Fig. 3B** and **3C**). Thus, regardless of their distinct genomic alterations, 5FU-induced MAPK signaling pathways in 5FU-resistant cells converges on phosphorylation of MEK1 either via PAK alone or both PAK and A-Raf (**Fig. 3M** and **3N**).

### 5FU stimulates divergent signaling cascades in 5FU-resistant GC cells

To support the results of the RPPA-based analysis, we also used GSEA for pathway profiling of 45FU and 74FU cells (**Fig. 4A–4D**). Unexpectedly, “G2/M checkpoint” was the top upregulated pathway in 45FU cells, but was the top downregulated pathway in 74FU cells, and vice versa for “IFNα response” (**Fig. 4A** and **4C**). To visualize the distinct signaling outcomes, we analyzed RPPA datasets focused on DDR for activation of “G2/M checkpoint” and STAT for “IFNα response” (**Fig. 4E–4H**). Although the DDR pathway is known to be upregulated upon 5FU treatment, to our knowledge, established 5FU-responsive regulators in the STAT pathway have not been reported. The DNA sensing cGAS-STING pathway is known to activate IRF3 and NFκB in the presence of damaged DNA^11^. Both IRF3 and transcriptional targets of NFκB like IRF1 and IRF7, can upregulate type I IFNs expression that in turn activates the STAT pathway. Thus, we included proteins and phosphoproteins in the NFκB pathway to assess potential IFNα responses. Consistent with the GSEA data, 45FU cells showed increased DDR pathway activity in response to 5FU, as indicated by higher amplitudes of ATR-pS^428^, p53-S^15^, and p21 expression compared to their parental cells, although levels of ATR-pS^428^ were high at baseline (**Fig. 4E**). Of note, 45FU cells are characterized by decrease in cleaved PARP levels, suggesting that intact PARP could detect DNA lesions and recruit repair proteins to the DNA damage^37^. In contrast to 45FU cells, 74FU cells exhibit little or no changes in DDR pathway protein levels except for increased levels of cleaved caspases (**Fig. 4F**). As indicated by increased levels of myeloid differentiation factor 88 (MYD88), TAK1-binding protein 2 (TAB2)-pS^372^ and IκBα-pS^32/36^, the NFκB pathway was activated by 5FU in 45FU cells (**Fig. 4G**). Meanwhile, 74FU cells display decreased levels of MYD88 and TAB2-pS^372^ at baseline (**Fig. 4H**), and instead have higher amplitudes of STAT1-pY^701^ and STAT3-pS^727^ expression compared to 45FU cells (**Fig. 4G** and **4H**). This result is in good agreement with GSEA data showing “IFNα response” and “IFNγ response” among the most upregulated pathways (**Fig. 4C**).

**Figure 4.**
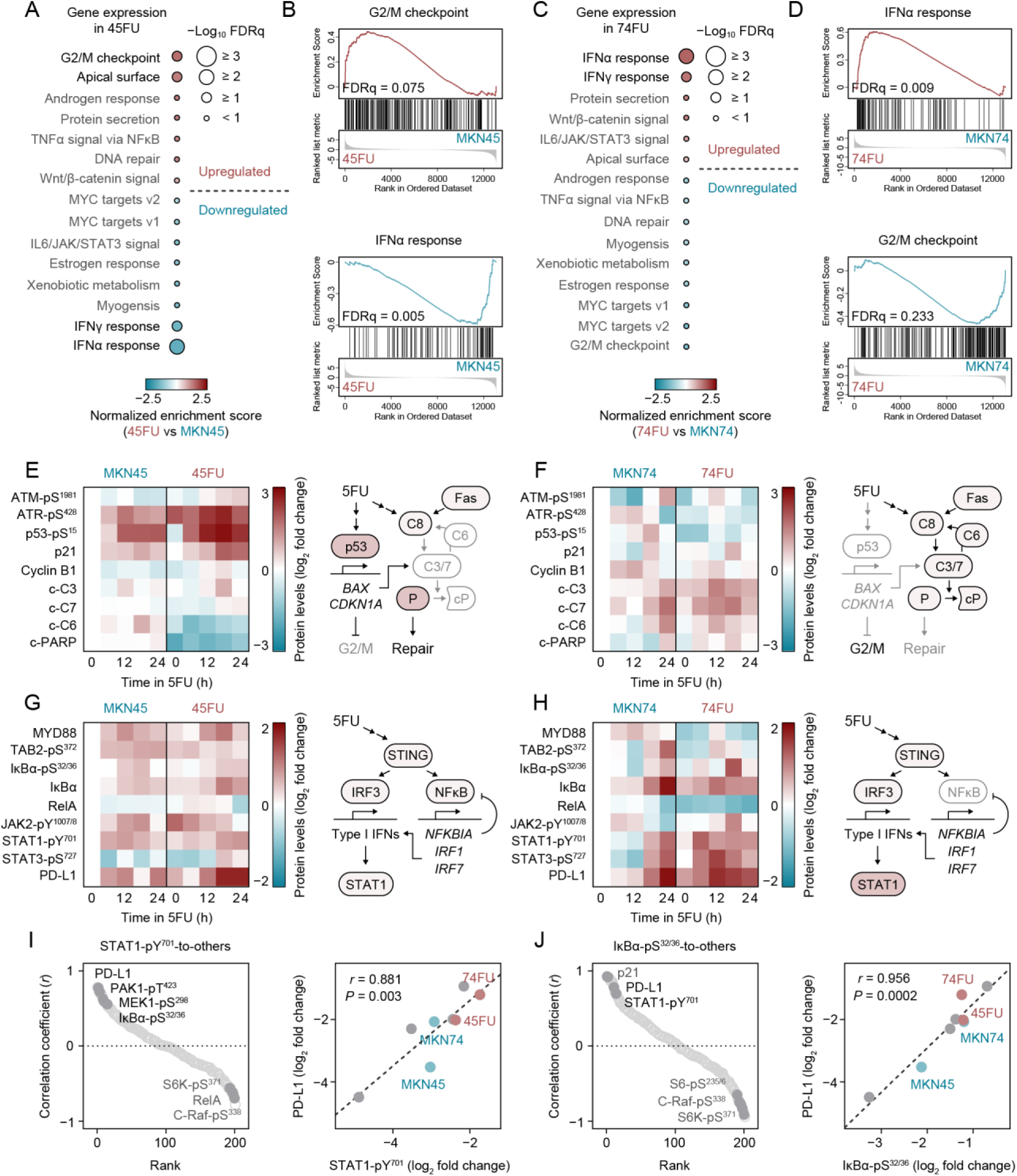
5FU elicits PD-L1 expression in 5FU-resistant GC cells. (**A**) Hallmark GSEA signatures from RNA-seq data ranked by NES for 45FU vs. parental MKN45 cells. (**B**) GSEA plots showing positive and negative enrichment of “G2/M checkpoint” and “IFNα response” gene sets in 45FU cells. (**C**) Hallmark GSEA signatures from RNA-seq data ranked by NES for 74FU vs. parental MKN74 cells. (**D**) GSEA plots showing positive and negative enrichment of “IFNα response” and “G2/M checkpoint” gene sets in 74FU cells. (**E**) DNA damage response (DDR) pathway-focused RPPA analysis of 45FU and parental MKN45 cells (left) and a schematic summarizing signaling outcomes in 45FU cells (right). (**F**) DDR pathway-focused RPPA analysis of 74FU and parental MKN74 cells (left) and a schematic summarizing signaling outcomes in 74FU cells (right). c-PARP, cleaved PARP; c-C3−9, cleaved caspase-3−9. (**G**) STAT and NFκB (proinflammatory) pathway-focused RPPA analysis of 45FU and parental MKN45 cells (left) and a schematic summarizing signaling outcomes in 45FU cells (right). (**H**) Proinflammatory pathway-focused RPPA analysis of 74FU and parental MKN74 cells (left) and a schematic summarizing signaling outcomes in 74FU cells (right). (**I**) Spearman’s rank correlation analysis to determine the correlation between STAT1-pY^701^ expression and expression of other proteins (left) and a representative scatter plot showing correlation between STAT1-pY^701^ and PD-L1 (right). (**J**) Spearman’s rank correlation analysis to determine the correlation between IκBα-pS^32/36^ expression and expression of other proteins (left) and a representative scatter plot showing correlation between IκBα-pS^32/36^ and PD-L1 (right). FDRq, false discovery rate (*q* value) (**B** and **D**).

### 5FU specifically elicits PD-L1 expression via STAT1 phosphorylation

Notably, both 45FU and 74FU cells demonstrated increased expression of programmed death ligand-1 (PD-L1) in response to 5FU (**Fig. 4G** and **4H**). A previous study reported that surface expression of PD-L1 and phosphorylation of STAT1 in pancreatic cancer cell lines was elicited by treatment with conventional chemotherapeutic drugs including 5FU, gemcitabine, and paclitaxel, although the correlation with phenotypic drug response was not clear^38^. To clarify whether non-DNA damaging drugs like paclitaxel and DTX have similar effects as DNA damaging drugs on IFN responses, we used GSEA to compare expression of IFN-responsive genes in GC cell lines with and without CIS, ETP, 5FU, or DTX treatment. Interestingly, expression of IFNα and IFNγ pathway genes was upregulated by treatment with CIS, ETP, or 5FU, but not with DTX, whereas NFκB pathway genes were upregulated by all drugs tested (**Supplementary Fig. 4**). To examine the significance of PD-L1 expression among the proteins that are coexpressed with STAT1-pY^701^, we determined the correlation between expression levels of STAT1-pY^701^ and the other proteins or phosphoproteins using Spearman’s rank correlation analyses (**Fig. 4I**). In line with the scenario, PD-L1 correlation is ranked on top along with PAK1-pT^423^/PAK2-pT^402^, MEK1-pS^298^, and IκBα-pS^32/36^. The correlation between expression levels of IκBα-pS^32/36^ and these other proteins further emphasized an association between STAT1 and PD-L1 (**Fig. 4J**). Collectively, these results suggest that 5FU elicits not only divergent signaling cascades in 5FU-resistant cells but also convergent PD-L1 expression.

### Lymphocyte count predicts a survival benefit with adjuvant chemotherapy

PD-L1, also known as B7-H1 or CD274, is a ligand for programmed cell death-1 (PD-1) that inhibits T cell activation by binding to PD-1 on T cells^39^. Anti-PD-L1/PD-1 antibodies together with an S-1 adjuvant chemotherapy containing the 5FU prodrug tegafur^3^, has become a standard treatment for locally advanced GC in Asia^40^. Based on the observation that the two 5FU-resistant cell lines showed increased levels of PD-L1 compared to their parental cell lines, we examined whether PD-L1 could be a prognostic factor for GC patients who underwent gastrectomy followed by S-1 adjuvant chemotherapy using the Northern Japan Gastric Cancer Study patient cohort^41,42,43^. Unexpectedly, neither overall survival (OS) nor the relapse-free survival (RFS) rate differed between surgery and S-1 adjuvant chemotherapy in PD-L1^−^ patients (**Fig. 5A**; **Supplementary Fig. 5A**). The most remarkable effect was seen when patients were segregated into two groups based on median total lymphocyte count (TLC), where S-1 for TLC^+^ patients demonstrated higher 5-year OS and RFS rates in GC patients than that of surgery alone (5-year OS: 80.7% vs. 67.2%; hazard ratio: 0.46; 95% confidence interval, 0.23‒0.88; *P* = 0.020; 5-year RFS: 76.5% vs. 63.3%; hazard ratio: 0.49; 95% confidence interval, 0.26‒0.8.9; *P* = 0.020) (**Fig. 5B** and **5C**; **Supplementary Fig. 5B**–**5D**). These observations indicate that host could contribute to relapse after S-1 adjuvant chemotherapy in addition to tumor cell-intrinsic 5FU-resistance or immunosuppression mechanisms (i.e., activation of PAK-MEK1 pathways or PD-L1 expression). In line with this scenario, *H. pylori* infection, which can recruit T lymphocytes and macrophages to gastric mucosa^44^, might confer a potential survival benefit in patients with advanced GC even after gastrectomy^45^. However, despite a strong correlation with inflammation scores for gastric mucosa based on the Sydney System^46^, the presence of *H. pylori* does not seem to be directly related to TLC (**Fig. 5D** and **5E**; **Supplementary Fig. 5E** and **5F**), suggesting that TLC may reflect an event that is distinct from that of *H. pylori*-induced immunity. Whether *H. pylori*-induced or not, lymphocytes can nonetheless secrete antitumor cytokines such as TNFα/β, and IFNγ^47^. To examine whether lymphocyte-derived cytokines are associated with survival of patients with GC, we compared the hazard ratios for patient survival outcomes based on expression levels of genes regulated by NFκB and STAT1, which can be activated by TNFα/β and IFNγ signaling inputs, with an independent cohort^18^. Remarkably, expression levels of NFκB/STAT1 targets including *IRF1*, *CD274*, *IRF9*, and *STAT1* are associated with better OS in the cohort (**Fig. 5F** and **5G**). These expression levels are in good agreement with the correlation between TLC and survival outcomes in our cohort. In contrast, other NFκB targets such as *NFKBIA*, *IRF7*, and *IL6* appear to drive opposite functions and outcomes.

**Figure 5.**
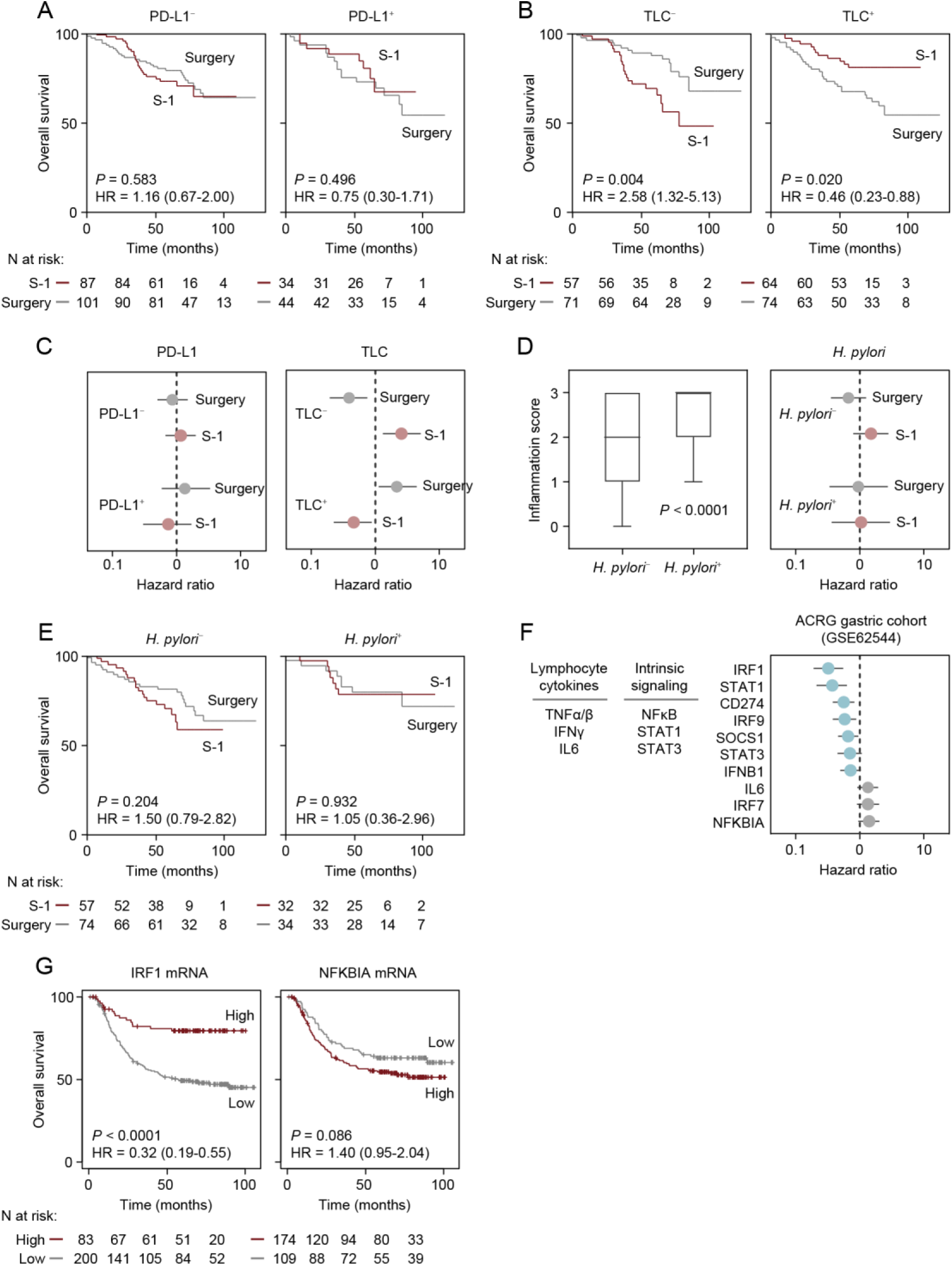
Survival curves for patients with stage II/III GC stratified by potential confounding factors. (**A**) Kaplan-Meier curves for overall survival (OS) in PD-L1^−^ (left) or PD-L1^+^ (right) patients treated with or without S-1. (**B**) Kaplan-Meier curves for OS in patients with low total lymphocyte count (TLC^−^) or high total lymphocyte count (TLC^+^) who did or did not receive S-1 treatment. (**C**) Subgroup analysis of OS in PD-L1^−^ (*n* = 188) or PD-L1^+^ (*n* = 78) patients (left) and TLC^−^ (*n* = 128) or TLC^+^ (*n* = 138) patients (right) evaluated for surgery alone and surgery followed by S-1 adjuvant chemotherapy. (**D**) Positive correlation between inflammation score and *H. pylori* positivity (left). Two-tailed *P* values were obtained with the nonparametric Mann-Whitney *U* test. Subgroup analysis of OS in *H. pylori*^−^ (*n* = 131) or *H. pylori*^+^ (*n* = 66) patients evaluated for surgery alone and surgery followed by S-1 adjuvant chemotherapy (right). (**E**) Kaplan-Meier curves for OS in *H. pylori*^−^ (left) or *H. pylori* ^+^ (right) patients who did or did not receive S-1. (**F**) Hazards for OS were evaluated for the expression of genes regulated by GC cell-intrinsic signaling in response to lymphocyte cytokines. (**G**) Kaplan-Meier curves for OS in ACRG GC patients (*n* = 283) stratified based on tumor *IRF1* (left) and *NFKBIA* (right) expression levels. Cox proportional hazards model was used to determine the hazard ratio of each mRNA expression level (**C**, **D**, and **F**). Error bars represent 95% confidence intervals. The *P* values were obtained with log-rank test (**A**, **B**, **E**, and **G**). Stratification strategies are shown in Supplementary Fig. 5G. ACRG, Asian Cancer Research Group^18^.

We next validated the finding that *NFKBIA* expression was associated with poor survival in patients with GC in an immunohistochemical study in our cohort and further examined whether IκBα expression could be a surrogate for low TLC (**Supplementary Fig. 6**). The number of patients who are TLC^−^ was significantly lower in IκBα^+^ patients than IκBα^−^ patients (**Supplementary Fig. 5G**). By excluding IκBα^−^ patients, S-1 for TLC^+^ patients demonstrated higher 5-year OS and RFS rates than for patients with GC who had surgery alone (5-year OS: 89.9% vs. 70.6%; hazard ratio: 0.26; 95% confidence interval, 0.06‒0.82; *P* = 0.026; 5-year RFS: 90.0% vs. 67.7%; hazard ratio: 0.22; 95% confidence interval, 0.05‒0.65; *P* = 0.008) (**Fig. 6A– 6D**). Exclusion of IκBα^−^ patients enabled more accurate outcome prediction compared to the method based on only TLC status (**Fig. 5B**; **Supplementary Fig. 5B**). Subgroup analysis showed the potential for substantial reduction in the risk of GC-related death and relapse after S-1 adjuvant chemotherapy in patients who were TLC-IκBα double-negative (**Fig. 6C** and **6E**). In addition, S-1 adjuvant chemotherapy provided no survival benefits for IκBα^+^ patients and notably the survival rate was even worse than surgery alone for TLC^−^ patients (**Fig. 6F–6J**).

**Figure 6.**
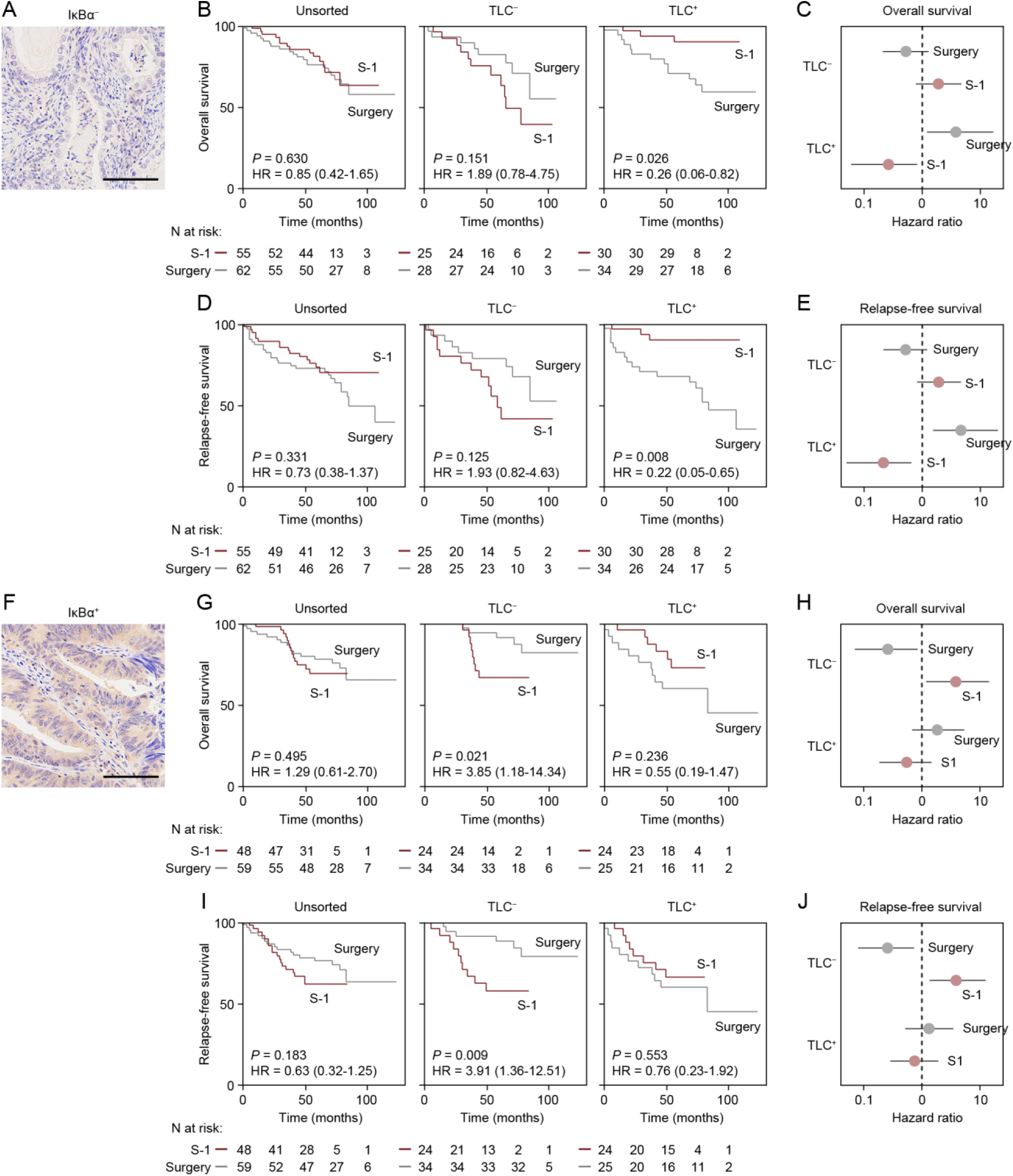
Survival curves for patients with stage II/III GC stratified by IκBα level and potential confounding factors. (**A**) Representative immunohistochemical staining in a IκBα^−^ tumor region. (**B**) Kaplan-Meier curves for OS in IκBα^−^ patients stratified by treatment (i.e., S-1 and Surgery) divided into TLC^−^ and TLC^+^ groups. (**C**) Subgroup analysis stratified by interaction of TLC^−^ (*n* = 53) or TLC^+^ (*n* = 64) based on the hazard for OS was evaluated by treatment. **(D)** Kaplan-Meier curves for RFS in IκBα^−^ patients stratified by treatment divided into TLC^−^ and TLC^+^ groups. **(E)** Subgroup analysis stratified by interaction of TLC^−^ (*n* = 53) or TLC^+^ (*n* = 64) based on the hazard for OS was evaluated by treatment. (**F**) Representative immunohistochemical staining in a IκBα^+^ tumor region. (**G**) Kaplan-Meier curves for OS in IκBα^+^ patients stratified by the treatment divided into TLC^−^ and TLC^+^ groups. (**H**) Subgroup analysis stratified by interaction of TLC^−^ (*n* = 58) or TLC^+^ (*n* = 49) based on the hazard for OS was evaluated by treatment. (**I**) Kaplan-Meier curves for RFS in IκBα^+^ patients stratified by treatment divided into TLC^−^ and TLC^+^ groups. (**J**) Subgroup analysis stratified by interaction of TLC^−^ (*n* = 58) or TLC^+^ (*n* = 49) based on the hazard for RFS was evaluated by treatment. Scale bar, 50 μm (**A** and **F**). Risk for survival was evaluated using Cox proportional hazards models (**C, E, H** and **J**). Error bars represent 95% confidence intervals. The *P* values were obtained with a log-rank test (**B, D, G** and **I**).

Collectively, TLC predicts a survival benefit for patients with advanced GC who received postoperative S-1 adjuvant chemotherapy. Thus, drug-resistance mechanisms involving NFκB signaling may allow GC cells to respond to lymphocyte-derived cytokines such as TNFα/β and IFNγ in the patients’ body.

## Discussion

TCGA and ACRG established a robust molecular classification method for GCs based on their mutations and gene expression profiles^17,18^. This classification method has been successfully applied to cell line-based compound screening for targeting the EMT subtype^48^. In this study, we classified eight GC cell lines into drug-sensitive and -resistant groups based on GI_50_ profiles in addition to comparing matched resistant-parental pairs to determine whether the phenotypic drug response was associated with known GC subtypes. Based on an apparent correlation with vimentin expression, cell lines having dual CIS/ETP-sensitivity are considered to be an EMT subtype (i.e., vimentin^+^), into which 15% of gastric tumors are classified^18^. Dual CIS/ETP sensitivity was also correlated with CNV represented by copy number loss of proximal 4q12–q13. Consistent with these observations, cell lines with dual CIS/ETP-sensitivity have decreased expression of DNA repair genes. These cell lines instead possess a highly sensitive caspase activation cascade towards caspase-3/7 that gives a sharp rise in PARP cleavage in the presence of CIS or ETP. Conversely, dual CIS/ETP-resistant cell lines exhibited upregulated expression of DNA repair genes and balanced caspase-3/7 activity whether or not CIS or ETP is present. For therapeutic purposes, nicotinamide phosphoribosyltransferase (NAMPT) inhibitors like FK866 and KPT9274 that are reported to selectively target the EMT subtype^48,49^ could be a substitute for CIS, especially when combined with 5FU, for patients with advanced GC. CIS-resistant GC cells could potentially be eliminated by PARP inhibitors such as olaparib, rucaparib, and niraparib based on their PARP dependency, and thus sequential administration of 5FU/CIS and PARP inhibitors warrants evaluation clinically^50^.

In contrast to CIS/ETP, 5FU sensitivity had no apparent correlation with known GC subtypes. Our exploratory pathway analysis based on protein expression dynamics after drug administration revealed that 5FU-resistant cell lines, but not cell lines that are resistant to other drugs, have diminished global protein dynamics compared to their sensitive counterparts, with the exception of the MAPK pathway. Subsequent pairwise comparisons of matched 5FU resistant-parental cell line pairs identified PAK-mediated phosphorylation of MEK1 at S298. MEK1 was the only MAPK component that remained active in the presence of 5FU in both 45FU and 74FU cells. In addition to MEK1 phosphorylation, both 5FU-resistant cell lines exhibited increased levels of PD-L1, regardless of the predominant signaling pathways (i.e., G2/M checkpoint activation in 45FU and increased IFN response in 74FU). Interestingly, high levels of PD-L1 are associated with chronic DNA damage in human epithelial cancer cells^51^, which could also be mimicked by long-term 5FU culture under which the 45FU and 74FU cell lines arose^15^.

Based on the finding that high PD-L1 expression was shared by both 5FU-resistant GC cell lines despite their distinct molecular profiles, we investigated the relationship between PD-L1 expression and prognosis in our cohort of patients with advanced GC. Despite its immunosuppressive function, PD-L1 expression was associated with a lower risk of relapse in patients with advanced GC who underwent gastrectomy followed by S-1 adjuvant chemotherapy. Among the potential confounding factors we investigated, TLC was the most significantly associated with lower risk of GC relapse, suggesting that host immunity could be a major contributor to patient survival. A similar phenomenon was seen for the classical NFκB-dependent survival mechanism in GC. Nuclear expression of RelA is positively correlated with overall survival rate of patients with GC^52^. More recent work showed that IRF1, a downstream target of NFκB, opposes multidrug resistance in GC cells^53^, although the involvement of NFκB or host lymphocytes has not been adequately explored. In addition to IRF1, we found that gene expression of STAT1, SOCS1, and PD-L1, which can be induced by Janus kinase (JAK)-STAT signal transduction induced by lymphocyte-secreted IFNγ, was also associated with favorable prognosis for patients with GC. These observations suggest that cytokines produced by lymphocytes such as TNFα/β and IFNγ are key components that could be exploited to prevent GC progression. Finally, based on the findings that *NFKBIA* expression was associated with poor survival in patients with GC, we examined whether expression of IκBα encoded by *NFKBIA* could be an indicator of poor outcomes in GC patients in our validation cohort. Strikingly, IκBα stratification could identify TLC^+^ GC patients do not have a survival benefit from postoperative chemotherapy.

The present study does have some limitations. First, we used only cell lines for quantitative molecular/phenotypic assays without morphological/localization information. Findings for stress responses and inflammation may require imaging evaluation as well as an assay to clarify the host-tumor response. Second, we did not treat the cell lines with simultaneous and/or pulse drug treatment, which is often applied in daily clinical practice. Third, the studies did not contain a functional immune response or other components of the tumor microenvironment that may be directly affected by the DNA-damaging agents as well as by bidirectional communication with the malignant cells. Finally, the clinical efficacy of potential molecular targeting agents after treatment with DNA-damaging agents remains to be clarified.

Collectively, our comprehensive molecular profiling of GC cell lines, including 5FU-responsive phosphoprotein dynamics, indicates that the mechanisms of 5FU resistance observed in cell line models may be overcome by host immunity. These studies suggest further determination of the best strategies for treatment of advanced CG patients.

## Methods

### Cell culture

GCIY, GSS, MKN1, MKN45, and MKN74 cells were obtained from the RIKEN Cell Bank. 5FU-resistant lines 45FU and 74FU were established from MKN45 and MKN74 cells, respectively, as described previously^15^. IWT-1 is a cell line that was established in our laboratory from a male Japanese patient with GC who had relapsed with peritonitis carcinomatosa^54^. Use of the IWT-1 cell line was approved by the Iwate Medical University Institutional Review Board (H25-116, and HG H25-15), and written informed consent was obtained from the family of donor patient, who had died at the time the cell line was established. The IWT-1 cell line is now distributed via the RIKEN BRC Cell Bank (https://cell.brc.riken.jp/en/). All eight cell lines were grown in RPMI 1640 media (Life Technologies, Carlsbad, CA, USA) supplemented with 10% fetal bovine serum (FBS) (Life Technologies), and cultured at 37°C in a humidified incubator supplied with 5% CO_2_.

### Drugs

5FU and DTX were purchased from Kyowa-Hakko Bio (Tokyo, Japan), and Sanofi KK (Tokyo, Japan), respectively. CIS and ETP were purchased from Nippon Kayaku (Tokyo, Japan).

### Growth suppression assay

Cells were plated in a 96-well plate at 1.0 to 4.0 × 10^4^ cells/well. Cell viability was determined using CCK-8 (Dojindo, Kumamoto, Japan) and a TriStar LB 941 microplate reader (Berthold Technologies, Bad Wildbad, Germany). GI_50_ values were calculated using GraphPad Prism software version 7.0 (GraphPad software Inc., San Diego, CA, USA).

### Targeted gene sequencing

Genotyping using the Ion AmpliSeq Pharmacogenomics Research Panel was conducted according to the manufacturer’s instructions using the Ion Torrent platform (Thermo Fisher Scientific, Waltham, MA, USA). This panel analyzes genetic variants including SNPs, insertion/deletions, and CNVs that are associated with 40 drug metabolizing enzymes. Gene mutation profiling was also performed using the Ion AmpliSeq Comprehensive Cancer Panel that covers nearly all coding regions of 409 genes that are known to be related to cancer (Thermo Fisher Scientific).

### RNA-seq

Total RNA was extracted using an RNeasy Kit (Qiagen), according to the manufacturer’s instructions. RNA-Seq libraries were constructed using TruSeq Stranded mRNA HT Sample Prep Kit (Illumina Inc, San Diego, CA, USA). Sequencing was carried out on the Illumina HiSeq3000 platform with a 36-base-single end run. Quality control of RNA-seq reads was performed using FastQC version 0.11.5. Raw sequence reads were mapped to the UCSC genome for human (hg19) using HISAT2 version 2.1.0, without novel splice variant discovery. Fragments per kilobase of exon per million (FPKM) mapped reads were calculated from the mapped reads using Ballgown version 2.6.0. FPKM data was later used in gene set enrichment analysis (GSEA) performed with GSEA Software (version 4.1.0) and WebGestalt^55^.

### qRT-PCR

Total RNA was extracted from GC cell lines using an RNeasy kit (QIAGEN). cDNA was synthesized from 500 ng total RNA in a 10 μl reaction volume using PrimeScript RT Master Mix (TaKaRa Bio, Otsu, Japan). Quantitative RT-PCR (qRT-PCR) was performed using the LightCycler Nano System (Roche, Mannheim, Germany). Primer sequences are shown in Supplementary Table 4.

### Reverse-Phase Protein Array (RPPA)

For sample collection, cells were exposed to anticancer drugs (5FU, CIS, ETP, and DTX) in a 96-well plate at low, medium, and high concentrations (**Supplementary Table 2**). Each concentration was applied at five time points over 24 h. Cells were collected by trypsinization followed by pipetting and centrifugation at 1,700 g for 2 min at 4 °C. The resulting cell pellet was stored at - 80 °C until further RPPA analysis as previously described^56^. For RPPA assays, cell pellets were processed to obtain cell lysates according to previously published protocols^56^. An individual RPPA contains 2,000 dots for 12 conditions ×6 time courses ×8 cell lines, and 800 dots for control MIX samples^57,58^. The dots were printed on nitrocellulose-embedded glass slides (Grace BioLabs, Bend, OR, USA) with an Aushon 2470 Microarrayer (Quanterix, Billerica, MA, USA). The Mix sample contains a variety of sample lysates from all possible drug administration conditions. Each sample lysate was spotted as nine serial 2-fold dilutions in tetraplicate for quantitative analysis and subsequently probed with individual primary antibodies (**Supplementary Table 3**) for which the specificity was verified by strip Western blotting^57,58^. To block nonspecific interactions, SuperG (Grace Biolabs), iBlock (Life Technologies), or standard BSA solutions were used depending on the antibody. Signals were obtained using a TSA kit (Thermo Fisher Scientific) and subsequently quantified using a TissueScope 4000 Scanner (Huron Technologies, Waterloo, ON).

### Cohorts (Discovery and Validation)

GSE62254 data set (*n* = 300) was used to determine lymphocyte cytokine-responsive genes that are associated with survival of GC patients. The Northern Japan Gastric Cancer Study patient cohort (NCT01905969, *n* = 658) was used to examine the effects of TLC, tumor PD-L1 and IκBα expression, and *H. pylori* positivity on treatment outcomes in advanced GC patients.

### Immunohistochemistry

A tissue microarray (TMA) including 658 GC specimens was made for immunohistochemistry (IHC) staining. The primary antibodies used were: rabbit anti-IκBα polyclonal antibody (E-AB-70089, Elabscience, Houston, TX, USA), mouse anti-PD-L1 monoclonal antibody (22C3, Dako, Santa Clara, CA, USA), and CD4/CD8 (4B12/SP16; Biocare Medical, Concord, Pacheco, CA, USA). For IκBα scoring, cases in which >30% of all tumor cells were stained were defined as positive. The staining evaluation was focused on the cytoplasm in the epithelial component. For staining evaluation, the MKN1 cell line was used as a positive control, and cases having equal or greater positivity relative to the positive control were considered positive. All scoring was performed by an independent pathologist (A.Y-A.), who was blinded to the clinical outcomes. The scoring algorithms and staining evaluation for PD-L1 and CD4/CD8 have been described previously^42^.

### Statistics

Data are shown as mean ± s.d. unless otherwise stated. Statistical analysis was performed using GraphPad Prism (GraphPad Software). Statistical significance of differences between two groups was evaluated using an unpaired two-tailed *t* test, or as indicated in the figure legends. For the correlation analysis, Pearson correlation coefficient *r* and *P* value in linear regression were used to evaluate the strength and statistical significance of the correlation. Differences having a *P* value of < 0.05 were considered statistically significant for screening purposes. For heatmap visualization, genes or proteins were ordered by average-linkage clustering, complete-linkage clustering, or from highest to lowest values in a column, unless otherwise stated. For survival analysis, patients were divided into two groups based on the median lymphocyte count or IHC positivity.

## Data availability

Targeted gene sequencing and RNA sequencing data have been deposited in the DNA Data Bank of Japan (DDBJ) via the Bioinformation and DDBJ Center under data set identifier DRA011072 (http://www.ddbj.nig.ac.jp). RPPA data is available at The University of Texas MD Anderson Cancer Center RPPA Data Repository under data set identifier TCPA00000007 (https://tcpaportal.org). Cohort data for advanced GC patients from the Northern Japan Gastric Cancer Study Consortium are available upon request.

## Acknowledgments

The authors thank the members of the Northern Japan Gastric Cancer Study Consortium and the patients who participated in this study. This work was supported by a Grant-in-Aid for Young Scientists (B) for K.K. (JP16K18458); Scientific Research (C) for S.S.N. (JP25462034); and Scientific Research on Innovative Areas for S.S.N. (JP16H01578 and 16H06279) of JSPS KAKENHI. K.K. and S.S.N. were supported by grant No. Y118 and 131, respectively, from the Keiryokai Research Foundation. The RPPA Core at The University of Texas MD Anderson Cancer Center is supported by NCI Grant #CA16672, NHIR50 Grant #R50CA221675; Functional Proteomics by Reverse Phase Protein Array in Cancer.

## Author contributions

K.K. and S.S.N. conceived and designed the experiments; K.K., S.S.N., T.I., A.Y-A., Y.K., A.K., K.W., H.H., V.C., J.W., V.E., D.R.S., and Y.L. performed the experiments; K.K., M.I., K.T., Y.Suzuki, Y.Sasaki, T.T., and S.S.N. performed the data analysis; E.P.III., L.A.L., and G.B.M. contributed to project oversight and advisory roles; K.K. and S.S.N. wrote the manuscript with assistance, comments, and final approval from all authors.

## Competing interests

T.I.: Grant/Research Support, Nippon Kayaku, Chugai Pharmaceutical, Daiichi Sankyo, Taiho Pharmaceutical, Otsuka Pharmaceutical, Quantdetect Inc.; Consultation, Quantdetect Inc. A.Y-A.: Consultation, PCL Japan, Kotobiken Medical Laboratories, LSI Medience, Quantdetect Inc. H.H.: Grant/Research Support, LSI Medience Co.; Consultation, Quantdetect, Inc. J.W.: Consultation, Baylor College of Medicine; Stock/Options/Financial, Theralink Technologies Inc. V.E.: Consultation, TheraLink Technologies, Inc.; Royalty distributions from patents licensed from George Mason University. Y.Sasaki: Grant/Research Support from Eli Lilly Japan. E.P.III.: Grant/Research Support, Springworks Therapeutics, Inc., Deciphera Pharmaceuticals, Genentech/Roche, Inc; Consultation, Ceres Nanosciences, Inc., Theralink Technologies, Inc., Perthera, Inc.; Stock/Options/Financial, Ceres Nanosciences, Inc., Theralink Technologies, Inc., Perthera, Inc. G.B.M.: Grant/Research Support, AstraZeneca, Zentalis, Nanostring, Ionis (Provision of tool compounds); Scientific Advisory Board/Consultant, Amphista, Astex, AstraZeneca, Biodyne, BlueDot, Chrysallis Biotechnology, Ellipses Pharma, GSK, ImmunoMET, Infinity, Ionis, Leapfrog Bio, Lilly, Medacorp, Nanostring, Neophore, Nuvectis, Pangea, PDX Pharmaceuticals, Qureator, Roche, Rybodyne, Signalchem Lifesciences, Tarveda, Turbine, Zentalis Pharmaceuticals; Stock/Options/Financial, Bluedot, Biodyne, Catena Pharmaceuticals, ImmunoMet, Nuvectis, RyboDyne, SignalChem, Tarveda, Turbine; and Licensed Technologies, HRD assay to Myriad Genetics and DSP to Nanostring. S.S.N.: Grant/Research Support, Array Jet, Taiho Pharmaceuticals, Boehringer-Ingelheim, Geninus, and Thermo Fisher Scientific; Honorarium, Chugai Pharmaceuticals and MSD; Consultation, Mills Institute for Personalized Care (MIPC); Advisor, CLEA Japan; and CEO, Quantdetect, Inc. The other authors declare no competing interests.

## Notes

### Funding Statement

This work was supported by JSPS KAKENHI (JP16K18458, JP25462034, JP16H01578, and 16H06279); grant from the Keiryokai Research Foundation (Y118 and 131); NCI Grant (CA16672); and NHI Grant (R50CA221675).

### Author Declarations

The multi-center study protocol was approved by the Institutional Review Board (IRB) of Iwate Medical University (H24-132, HGH24-12, and HG2020-018), where all analysis was carried out. Ethical approval for conducting this study was given by all IRB committees of all 12 participating institutions listed in the Northern Japan Gastric Cancer Study Consortium (Nishizuka et al., J Surg Oncol, 2018; Ohmori et al., J Surg Oncol, 2019; Koizumi et al., J Natl Cancer Inst, 2022).

