## Supplementary for "Dynamic phospho-proteogenomic analysis of gastric cancer cells suggests host immunity provides survival benefit"

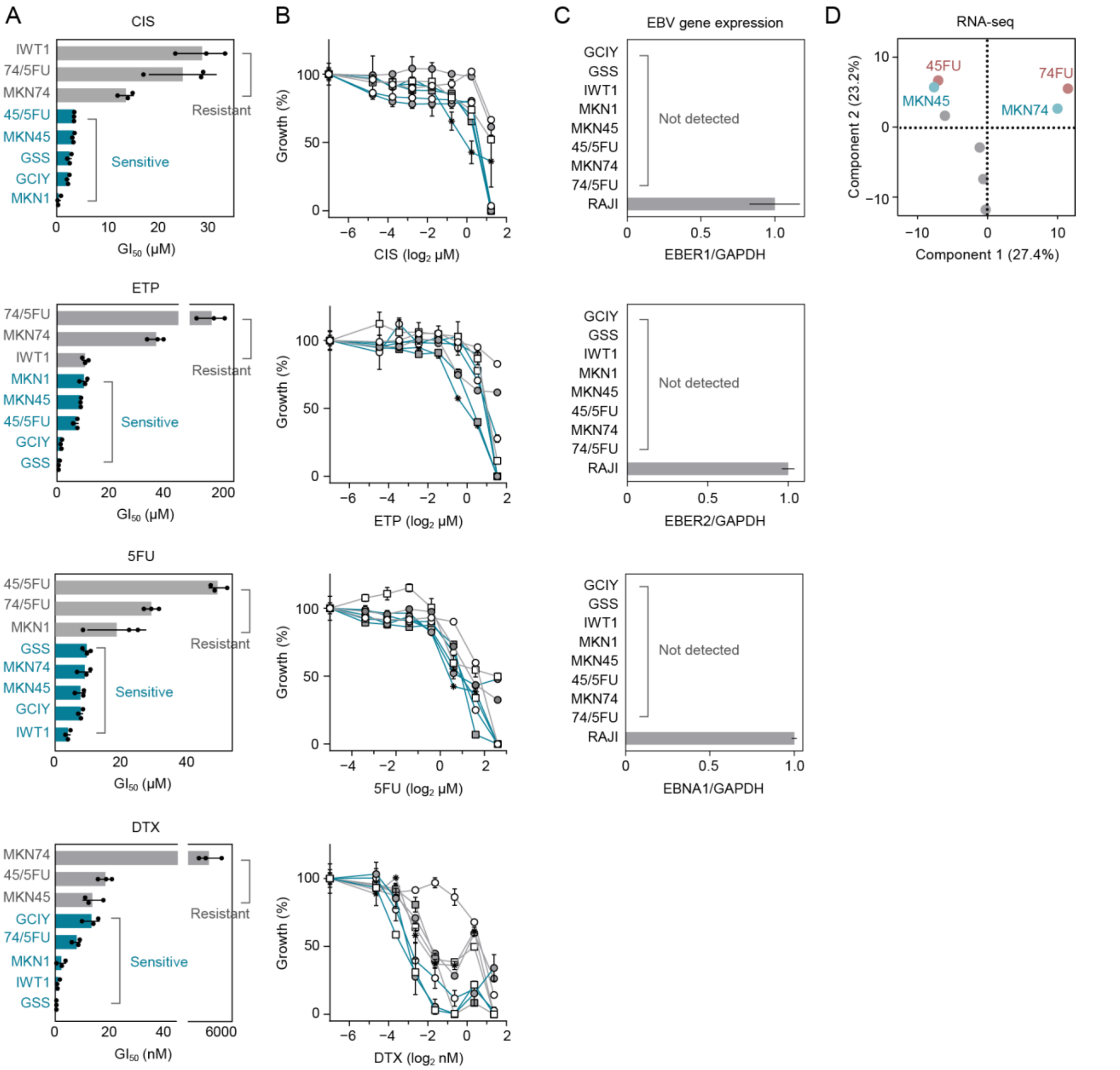


**Supplementary Figure 1.** Phenotypic and gene expression profiling of GC cell lines. (**A**) $GI_{50}$ profiles of GC cell lines to define sensitivity and resistance to each drug. (**B**) Dose-response curves for $GI_{50}$ by 5FU, CIS, ETP, and DTX. (**C**) Quantification of EBV gene expression in GC cell lines. GAPDH was used as an internal control. The EBV-positive cell line RAJI was used as a positive control. (**D**) Principal component analysis of RNA-seq gene expression data. Experiments were performed in triplicate (**A** and **B**) or duplicate (**C**). Error bars represent s.d. (**A**), s.e.m. (**B**) or range (**C**).

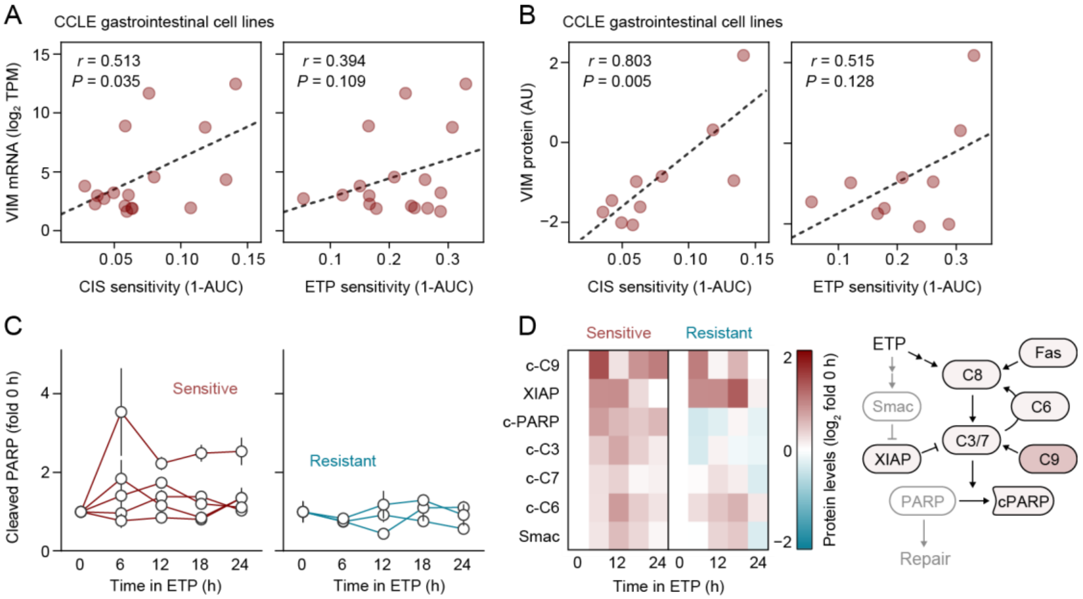


**Supplementary Figure 2.** ETP sensitivity associated with increased VIM expression and active PARP cleavage. (**A**) Scatter plots showing correlation between AUC values of CIS or ETP and *VIM* mRNA expression in CCLE gastrointestinal cancer cell lines. Each dot indicates an individual cell line. (**B**) Scatter plots showing correlation between AUC values of CIS or ETP and vimentin protein levels in gastrointestinal cancer cell lines. Each dot indicates an individual cell line. (**C**) Time course RPPA data showing changes in cleaved PARP levels after ETP treatment. Error bars represent s.e.m. (**D**) Caspase-focused RPPA analysis of dual CIS/ETP-sensitive (mean, $n$ = 5) and -resistant (mean, $n$ = 3) cell lines (left) and a schematic of ETP-activated signaling outcomes in dual CIS/ETP-sensitive cell lines (right). c-PARP, cleaved PARP; c-C3–9, cleaved caspase-3–9.

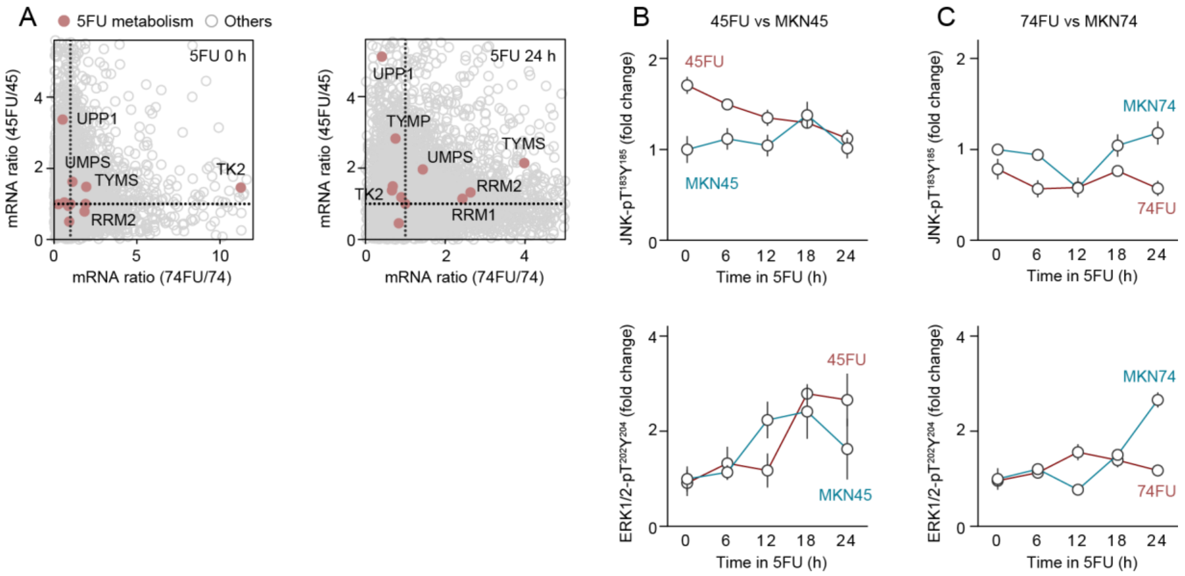


**Supplementary Figure 3.** Gene and protein expression profiling of 5FU-resistant cell lines. (**A**) Gene expression analysis of 5FU-resistant GC cell lines. mRNA levels in 45FU and 74FU are shown as fold-change compared to their matched parental cell lines and further highlighted for 5FU metabolism pathway genes. (**B**) Temporal changes in JNK-pT$^{183}$/Y$^{185}$, p38-pT$^{180}$/Y$^{182}$, and ERK1/2-pT$^{202}$/Y$^{204}$ levels in MKN45 and 45FU cell lines after 5FU treatment. (**C**) Temporal changes in JNK-pT$^{183}$/Y$^{185}$, p38-pT$^{180}$/Y$^{182}$, and ERK1/2-pT$^{202}$/Y$^{204}$ levels in MKN74 and 74FU cell lines after 5FU treatment. Error bars represent s.e.m.

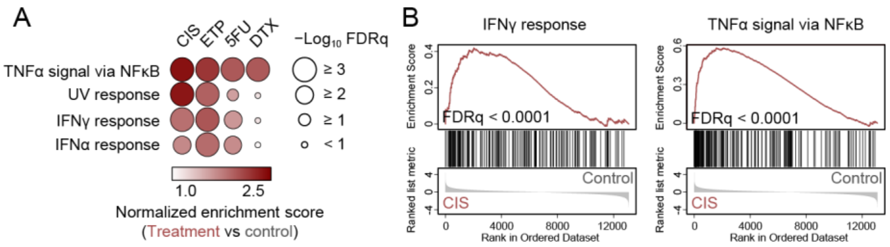


**Supplementary Figure 4.** Proinflammatory gene expression induced by chemotherapeutic drugs. (**A**) Enrichment of “TNFα signaling via NFκB”, “UV response”, “IFNγ response”, and “IFNα response” gene sets in GC cell lines ($n$ = 8) treated with the indicated drugs. “UV response” gene set was used as a positive control for DNA-damaging drugs. DTX was used as a non-DNA-damaging drug. (**B**) GSEA curves for “IFNγ response” and “TNFα signaling via NFκB”. FDRq, false discovery rate ($q$ value).

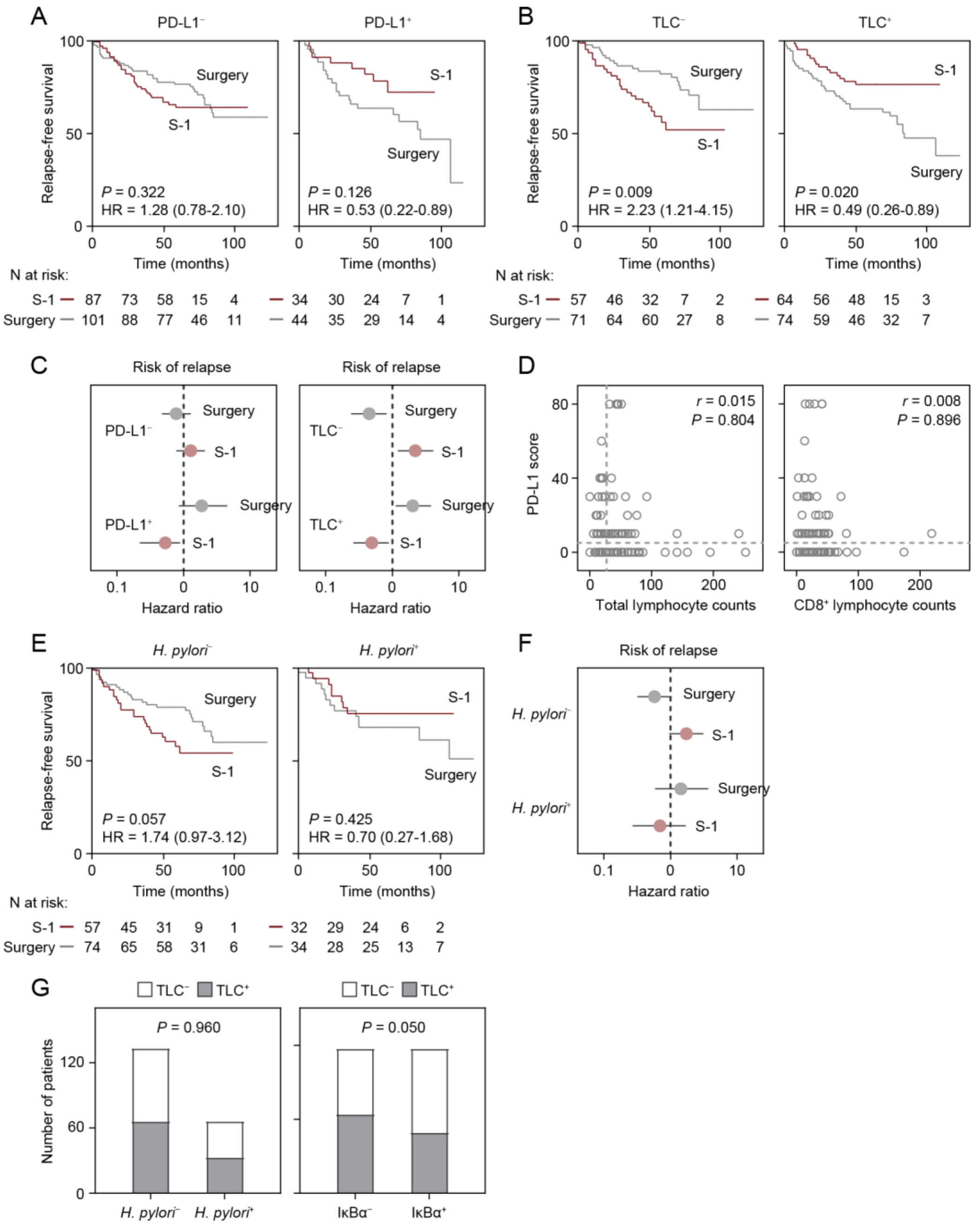


**Supplementary Figure 5.** RFS curves of stage II/III GC patients stratified by potential confounding factors. (**A**) Kaplan-Meier curves for relapse-free survival (RFS) in PD-L1$^{-}$ (left) or PD-L1$^{+}$ (right) patients by treatment (i.e., S-1 or Surgery). (**B**) Kaplan-Meier curves for RFS in patients with low total lymphocyte count (TLC$^{-}$) or high total lymphocyte count (TLC$^{+}$) by treatment. (**C**) Subgroup analysis stratified by interaction of PD-L1$^{-}$ (n = 188) or PD-L1$^{+}$ (n = 78);

and TLC$^{-}$ (n = 128) or TLC$^{+}$ (n = 138) based on the hazard for RFS was evaluated by treatment. (**D**) Scatter plots for PD-L1 score and total lymphocyte count (left) or CD8$^{+}$ lymphocyte count (right). (**E**) Kaplan-Meier curves for RFS in *H. pylori*$^{-}$ (left) or *H. pylori*$^{+}$ (right) patients by treatment. (**F**) Subgroup analysis stratified by interaction of *H. pylori*$^{-}$ (n = 131) or *H. pylori*$^{+}$ (n = 66) based on the hazard for RFS was evaluated by treatment. (**G**) Chi-square exact test for TLC and *H. pylori* positivity (left) and TLC and IκBα positivity (right). Risk of relapse was estimated using Cox proportional hazards models (**C** and **F**). Error bars represent 95% confidence intervals. The *P* values were obtained with log-rank test (**A**, **B**, and **E**).

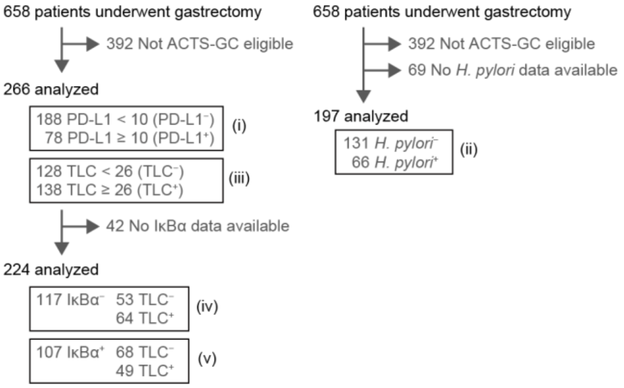


**Supplementary Figure 6.** Flowchart for study cohort selection. The number of patients in the stratified groups based on positivity for PD-L1 (i), *H. pylori* (ii), TLC (iii), and combined TLC and IκBα (iv and v) is shown. ACTS-GC, Adjuvant Chemotherapy Trial of S-1 for Gastric Cancer[4].

**Supplementary Table 1.** Gastric and colon cancer cell lines in the PRISM repurposing primary screen.

| Name | Primary disease | Depmap ID. |
|---|---|---|
| 2313287 | Gastric cancer | ACH-000948 |
| AGS | Gastric cancer | ACH-000880 |
| LMSU | Gastric cancer | ACH-000255 |
| MKN1 | Gastric cancer | ACH-000351 |
| MKN7 | Gastric cancer | ACH-000678 |
| NUGC3 | Gastric cancer | ACH-000911 |
| SH10TC | Gastric cancer | ACH-000764 |
| SNU668 | Gastric cancer | ACH-000344 |
| GP2D | Colon/Colorectal cancer | ACH-000982 |
| HCT116 | Colon/Colorectal cancer | ACH-000971 |
| HCT15 | Colon/Colorectal cancer | ACH-000997 |
| RCM1 | Colon/Colorectal cancer | ACH-000565 |
| RKO | Colon/Colorectal cancer | ACH-000943 |
| SNU81 | Colon/Colorectal cancer | ACH-000991 |
| SNUC2A | Colon/Colorectal cancer | ACH-000967 |
| SNUC4 | Colon/Colorectal cancer | ACH-000959 |
| SW48 | Colon/Colorectal cancer | ACH-000958 |

**Supplementary Table 2.** Drug concentrations for the time course RPPA analysis.

| Drug | Low | Middle | High |
|---|---|---|---|
| 5FU | 0.38 µM | 3.8 µM | 38 µM |
| CIS | 0.17 µM | 1.7 µM | 17 µM |
| ETP | 0.34 µM | 3.4 µM | 34 µM |
| DTX | 0.23 µM | 2.3 µM | 23 µM |

**Supplementary Table 3.** Primary antibodies used for RPPA.

| Target | Vendor | Catalog no. | Dilution |
|---|---|---|---|
| Abl SH2 domain | Sigma-Aldrich | 06-465 | 1:500 |
| AKT-pS$^{473}$ | Cell Signaling | 9271 | 1:100 |
| A-Raf-pS$^{299}$ | Cell Signaling | 4431 | 1:50 |
| ASK1-pS$^{83}$ | Cell Signaling | 3761 | 1:50 |
| ATF2-pT$^{71}$ | Santa Cruz | sc-8398 | 1:100 |
| ATF2-pT$^{71}$ | Cell Signaling | 9221 | 1:100 |
| ATG7 | Cell Signaling | 2631 | 1:100 |
| ATM-pS$^{1981}$ | Cell Signaling | 5883 | 1:100 |
| ATR-pS$^{428}$ | Cell Signaling | 2853 | 1:100 |
| Aurora A | Santa Cruz | sc-373856 | 1:100 |
| Aurora A-pT$^{288}$/B-pT$^{232}$/C-pT$^{198}$ | Cell Signaling | 2914 | 1:100 |
| BAD-pS$^{112}$ | Cell Signaling | 9291 | 1:200 |
| BAX | Cell Signaling | 2772 | 1:200 |
| Bcl2-pS$^{70}$ | Cell Signaling | 2827 | 1:100 |
| Bcl-xL | Cell Signaling | 2762 | 1:200 |
| β-actin | Santa Cruz | sc-47778 | 1:100 |
| β-arrestin1-pS$^{412}$ | Cell Signaling | 2416 | 1:100 |
| β-catenin | Cell Signaling | 9582 | 1:100 |
| β-catenin-pT$^{41}$/S$^{45}$ | Cell Signaling | 9565 | 1:100 |
| BIM | Cell Signaling | 2933 | 1:500 |
| Caspase-2 | BD | 611022 | 1:10 |
| Caspase-3 | Cell Signaling | 9662 | 1:100 |
| Caspase-3, cleaved | Cell Signaling | 9661 | 1:200 |
| Caspase-6, cleaved | Cell Signaling | 9761 | 1:50-100 |
| Caspase-7 | Cell Signaling | 9492 | 1:100 |
| Caspase-7, cleaved | Cell Signaling | 9491 | 1:100 |
| Caspase-9 | Cell Signaling | 9502 | 1:500 |
| Caspase-9, cleaved | Cell Signaling | 9505 | 1:100 |
| Caspase-9, cleaved | Cell Signaling | 9501 | 1:100 |
| CD13 | Santa Cruz | sc-13536 | 1:100 |
| CD133 | Cell Signaling | 9196 | 1:100 |
| CDK1 | BD | 61037 | 1:200 |
| CDK2 | BD | 61045 | 1:200 |
| CDK4 | BD | 61047 | 1:200 |
| Chk1-pS$^{345}$ | Cell Signaling | 2341 | 1:100 |
| c-kit/KIT | Santa Cruz | sc-365504 | 1:500 |
| c-kit/KIT | Thermo | 34-8800 | 1:500 |
| Cox-2 | BD | 610203 | 1:200 |
| CXCR4 | Santa Cruz | sc-53534 | 1:100 |
| Cyclin A | BD | 611268 | 1:50 |
| Cyclin A2 | Cell Signaling | 4656 | 1:50 |
| Cyclin B | BD | 610219 | 1:500 |
| Cyclin B1 | Cell Signaling | 4135 | 1:200 |
| Cyclin D1 | BD | 554180 | 1:100 |
| Cyclin D3 | Cell Signaling | 2936 | 1:500 |
| Cyclin E | Cell Signaling | 4129 | 1:500 |
| Cytokeratin 19 | Santa Cruz | sc-37126 | 1:1000 |
| Cytokeratin 8 | Santa Cruz | sc-8020 | 1:1000 |
| E-cadherin | Cell Signaling | 3195 | 1:50 |
| EGFR | Cell Signaling | 2232 | 1:100 |

| | | | |
|---|---|---|---|
| EGFR-pY$^{1045}$ | Cell Signaling | 2237 | 1:100 |
| ELK1-pS$^{383}$ | Cell Signaling | 9181 | 1:200 |
| EpCAM | Santa Cruz | sc-71057 | 1:100 |
| ErbB2/HER2 | Cell Signaling | 2242 | 1:100 |
| ERK1/2-pT$^{202}$/Y$^{204}$ | Cell Signaling | 9101 | 1:500 |
| ERK1/2 | Cell Signaling | 9102 | 1:100 |
| FADD-pS$^{194}$ | Cell Signaling | 2781 | 1:100 |
| GAPDH | Cell Signaling | 2118 | 1:500 |
| GSK3α | Cell Signaling | 9337 | 1:100 |
| GSK3α/β | Santa Cruz | sc-7291 | 1:100 |
| GSK3β | Cell Signaling | 9332 | 1:100 |
| HDAC1 | Cell Signaling | 2062 | 1:200 |
| HDAC3 | Cell Signaling | 2632 | 1:1000 |
| HDAC4 | Cell Signaling | 2072 | 1:100 |
| HDAC6 | Santa Cruz | sc-11420 | 1:2000 |
| Histone H3 | Abcam | ab1791 | 1:10000 |
| Histone H3-pS$^{28}$ | Sigma-Aldrich | 07-145 | 1:500-1000 |
| HMGB1 | Cell Signaling | 6893 | 1:200 |
| HSP70 | StressGen | SPA-810 | 1:200 |
| HSP90 | Cell Signaling | 4875 | 1:50-100 |
| HSP90-pT$^{5/7}$ | Cell Signaling | 3488 | 1:100-500 |
| IR-pY$^{1150/51}$/IGF1R-pY$^{1135/36}$ | Cell Signaling | 3024 | 1:500 |
| IGF1R β | Cell Signaling | 3027 | 1:1000 |
| IκBα | Cell Signaling | 9242 | 1:500-1000 |
| IκBα-pS$^{32/36}$ | Cell Signaling | 9246 | 1:100 |
| Jak2-pY$^{1007/1008}$ | Cell Signaling | 3771 | 1:200 |
| JNK/SAPK | BD | 610627 | 1:100 |
| JNK/SAPK | Cell Signaling | 9252 | 1:500 |
| JNK/SAPK-pT$^{183}$/Y$^{185}$ | Cell Signaling | 9251 | 1:100 |
| Ki67 | Cell Signaling | 4719 | 1:500 |
| LC3B | Cell Signaling | 2775 | 1:100 |
| LKB1-pS$^{428}$ | Cell Signaling | 3051 | 1:100 |
| MDR1 | Sigma-Aldrich | P7965 | 1:100 |
| MEK1/2 | Cell Signaling | 9122 | 1:1000 |
| MEK1/2-pS$^{217/221}$ | Cell Signaling | 9121 | 1:200 |
| MEK1-pS$^{298}$ | Cell Signaling | 9128 | 1:2000 |
| MGMT | Cell Signaling | 2739 | 1:100 |
| MLH1 | Cell Signaling | 3515 | 1:500 |
| Mst1-pT$^{183}$/Mst2-pT$^{180}$ | Cell Signaling | 3681 | 1:100 |
| mTOR | Cell Signaling | 2972 | 1:200 |
| mTOR-pS$^{448}$ | Cell Signaling | 2971 | 1:100 |
| MYD88 | Cell Signaling | 4283 | 1:500 |
| MYC | Santa Cruz | sc-40 | 1:100 |
| NANOG | Cell Signaling | 3580 | 1:1000 |
| NANOG | Cell Signaling | 4903 | 1:100 |
| N-cadherin | Cell Signaling | 4061 | 1:100 |
| NFκB/RelA | Cell Signaling | 3034 | 1:100 |
| Notch1 | Santa Cruz | sc-376403 | 1:100 |
| NPM-pT$^{199}$ | Cell Signaling | 3541 | 1:100 |
| NRF2 | Abcam | ab62352 | 1:200 |
| p21 | Santa Cruz | sc-6246 | 1:50-100 |
| p21 | Cell Signaling | 2946 | 1:200 |
| p27 | BD | 610242 | 1:100 |
| p27-pT$^{187}$ | Sigma-Aldrich | 71-7700 | 1:200 |

| | | | |
|---|---|---|---|
| p38 | Cell Signaling | 9212 | 1:200 |
| p38-p$T^{180}$/$T^{182}$ | Cell Signaling | 9211 | 1:100 |
| p53 | Invitrogen | AHO0152 | 1:200 |
| p53 | Thermo | MS-187 | 1:100 |
| p53-p$S^{15}$ | Cell Signaling | 9284 | 1:1000 |
| p70/p85 S6K | Cell Signaling | 2708 | 1:100-1000 |
| p70/p85 S6K | Cell Signaling | 9202 | 1:100-1000 |
| p70 S6K-p$S^{371}$ | Cell Signaling | 9208 | 1:50 |
| p70 S6K-p$T^{389}$ | Cell Signaling | 9234 | 1:500 |
| p90 RSK-p$T^{359}$/$S^{363}$ | Cell Signaling | 9344 | 1:200 |
| p90 RSK-p$S^{380}$ | Cell Signaling | 9341 | 1:200 |
| PAK1-p$S^{199/204}$/PAK2-p$S^{192/197}$ | Cell Signaling | 2605 | 1:100 |
| PAK1-p$T^{423}$/PAK2-p$T^{402}$ | Cell Signaling | 2601 | 1:100 |
| PARP, cleaved | Cell Signaling | 9541 | 1:100 |
| PCAF | Cell Signaling | 3378 | 1:500 |
| PDK1-p$S^{241}$ | Cell Signaling | 3061 | 1:200 |
| PD-L1 | Cell Signaling | 13684 | 1:5000 |
| PI3K p85 | Cell Signaling | 4292 | 1:500 |
| PI3K p85-p$Y^{458}$/p55-p$Y^{199}$ | Cell Signaling | 4228 | 1:100 |
| PIAS1 | Cell Signaling | 3550 | 1:100 |
| PKCα | Upstate | 05-154 | 1:100-200 |
| PKCβII-p$S^{660}$ (pan) | Cell Signaling | 9371 | 1:100 |
| PP2A-B | Cell Signaling | 4953 | 1:1000 |
| PRK1-p$T^{774}$/PRK2-p$T^{816}$ | Cell Signaling | 2611 | 1:100 |
| Proteasome 20S C2 | abcam | ab3325 | 1:500 |
| PTEN | Cell Signaling | 9552 | 1:50-100 |
| PTEN-p$S^{380}$ | Cell Signaling | 9551 | 1:500 |
| PUMA | Cell Signaling | 4976 | 1:200 |
| pY | Sigma-Aldrich | 05-321 | 1:1000 |
| pY | Cell Signaling | 9411 | 1:1500 |
| Raf1/C-Raf-p$S^{259}$ | Cell Signaling | 9421 | 1:100 |
| Raf1/C-Raf-p$S^{338}$ | Cell Signaling | 9427 | 1:200 |
| Ras (pan) | Santa Cruz | sc-166691 | 1:100-200 |
| Rb-p$S^{780}$ | Cell Signaling | 3590 | 1:2000 |
| RSK2 | Santa Cruz | sc-9986 | 1:200 |
| RSK3-p$T^{356}$/$S^{360}$ | Cell Signaling | 9348 | 1:500 |
| S6-p$S^{235/236}$ | Cell Signaling | 4858 | 1:200 |
| S6-p$S^{240/244}$ | Cell Signaling | 5364 | 1:200 |
| SEK1/MKK4-p$S^{80}$ | Cell Signaling | 9155 | 1:200 |
| Smac/Diablo | Cell Signaling | 2954 | 1:10000 |
| SMAD2-p$S^{465/467}$ | Cell Signaling | 3101 | 1:200 |
| SOX2 | Cell Signaling | 2748 | 1:100 |
| SSEA4 | Cell Signaling | 4755 | 1:100 |
| SRC | Santa Cruz | sc-18 | 1:200 |
| STAT1 | Cell Signaling | 9172 | 1:100-200 |
| STAT1-p$Y^{701}$ | Cell Signaling | 9171 | 1:500 |
| STAT3 | Cell Signaling | 9132 | 1:500 |
| STAT3-p$S^{727}$ | Cell Signaling | 9134 | 1:100 |
| SUMO1 | Cell Signaling | 4930 | 1:200 |
| Survivin | Cell Signaling | 2808 | 1:500 |
| TAB2-$S^{372}$ | Cell Signaling | 8155 | 1:100 |
| TRA-1-60 | Cell Signaling | 4746 | 1:100-200 |
| Tubulin | Cell Signaling | 2148 | 1:100-1000 |
| TWIST | Santa Cruz | sc-81417 | 1:100 |

| | | | |
|---|---|---|---|
| UBC3/Cdc34 | Cell Signaling | 4997 | 1:100 |
| Ubiquitin | Cell Signaling | 3936 | 1:50 |
| Ubiquitin (K48 linkage) | Cell Signaling | 4289 | 1:100 |
| VDR | Santa Cruz | sc-13133 | 1:100 |
| VHL | Cell Signaling | 2738 | 1:1000 |
| Wnt5a/b | Cell Signaling | 2530 | 1:100 |
| XIAP | Cell Signaling | 2042 | 1:100 |
| YAP-pS$^{127}$ | Cell Signaling | 13008 | 1:200 |
| ZO-1 | Cell Signaling | 5406 | 1:100 |

**Supplementary Table 4.** RT-PCR primers.

| Gene | Forward | Reverse |
|---|---|---|
| EBER1 | AGGACCTACGCTGCCCTAGA | GGGAAGACAACCACAGACAC |
| EBER2 | AGGACAGCCGTTGCCCTAGTGG | GCAAATGCTCTAGGCGGGAAG |
| EBNA1 | CCTCCCTGGTTTCCACCTAT | TCCTCACCCTCATCTCCATC |
| GAPDH | AATCCCATCACCATCTTCCA | TGGACTCCACGACGTACTCA |
